# Metformin use is associated with lower risks of dementia, anxiety and depression: The Hong Kong Diabetes Study

**DOI:** 10.1101/2024.01.07.24300938

**Authors:** Jeremy Man Ho Hui, Jiandong Zhou, Teddy Tai Loy Lee, Kyle Hui, Oscar Hou In Chou, Yan Hiu Athena Lee, Sharen Lee, Wing Tak Wong, Abraham Ka Chung Wai, Carlin Chang, Kamalan Jeevaratnam, Tong Liu, Gary Tse

## Abstract

**Aims:** To compare the effects of metformin and sulphonylurea on new-onset dementia, anxiety disorder and depression, and all-cause mortality in patients with type 2 diabetes mellitus.

**Methods:** This is a retrospective population-based cohort study of type 2 diabetes mellitus patients exposed to either metformin or sulphonylureas attending the Hospital Authority of Hong Kong between 1^st^ and 31^st^ December 2009. The follow-up was until 31^st^ December 2019. The primary outcome was a new diagnosis of dementia, and anxiety disorder/depression. Propensity score matching (1:1 ratio) between metformin and sulphonylurea users based on demographics, CAIDE score, CHA-DS-VASc score, Charlson comorbidity index, past comorbidities, medications, and total cholesterol was performed. Cox regression was used to identify significant risk predictors. Cause-specific and subdistribution hazard models were also used.

**Results:** A total of 89,711 patients (46% men, mean age: 67 years old [SD: 12]) followed-up for 1,579 days (SD: 650). Metformin users were at a lower risk of dementia (before: 0.78 [0.72, 0.84], P-value < 0.0001; after: 0.88 [0.80, 0.97], P-value = 0.0074), anxiety disorder and depression (before: 0.77 [0.69, 0.86], P-value < 0.0001; after: 0.71 [0.61, 0.82], P-value < 0.0001), and all-cause mortality (before: 0.69 [0.68, 0.71], P-value < 0.0001; after: 0.83 [0.80, 0.85], P-value < 0.0001). These associations remained significant in the competing risk models.

**Conclusion:** Metformin use is associated with lower risks of dementia, new-onset anxiety disorder and depression, and all-cause mortality, compared to sulphonylurea use. The protective effects of metformin and possible use in drug repurposing for indications beyond diabetes warrant further investigation.

**Highlights:**

- Patients with type 2 diabetes have an increased risk of cognitive, anxiety or depressive problems
- Metformin use was associated with lower risks of new diagnosis of dementia, anxiety disorder and depression
- Patients who developed dementia had lower levels of albumin, alanine transaminase and HbA1c compared to those who developed anxiety disorder and depression
- Appropriate glycemic control and maintenance of normal liver function are important in slowing cognitive decline in type 2 diabetes mellitus

## 1. Introduction

The prevalence of type 2 diabetes mellitus has been increasing globally, with approximately 462 million of the world’s population affected in 2017 (Khan et al., 2020). The disease can cause various complications, such as cardiovascular and neurological problems. Evidence from different studies show that patients with type 2 diabetes have an increased risk of cognitive dysfunction, such as dementia and depression (van Sloten et al., 2020; Whitmer, 2007). In patients with type 2 diabetes, reduced cognitive function might be associated with reduced self-care and increased use of care services, thus imposing an even greater burden on the healthcare system. There have been inconsistent findings in regard to how certain antidiabetic drugs influence the risk of developing cognitive dysfunction (Zhang et al., 2020). Furthermore, while it is established that type 2 diabetes is associated with an increased risk of dementia (Cheng et al., 2012), few recommendations are available to guide clinicians on how to approach cognitive dysfunction in patients with diabetes (Srikanth et al., 2020). It was suggested that medication control of type 2 diabetes would improve cognitive function among patients.

Metformin and sulfonylurea are the two commonest oral medications for type 2 diabetes, given that they are safe-to-use and well-tolerated (MacCallum and Senior, 2019; Sola et al., 2015). Thus, metformin and sulfonylurea have been used for decades to control blood glucose levels in patients with diabetes. This glycemic control has been shown to reduce micro- and macrovascular complications (Boussageon et al., 2011). Interestingly, metformin use has been associated with beneficial effects on the cardiovascular system (Lee et al., 2022a; Zhou et al., 2022) and may be protective against development of chronic diseases (Lee et al., 2022b), or slow disease progression in special populations such as cancer (Lee et al., 2023b). Although dementia and anxiety disorders are not classically described as microvascular complications of type 2 diabetes, there is growing evidence that these cognitive disorders are caused by microvascular dysfunction (De Silva and Faraci, 2016; Sierra, 2012). Since the pathogenesis of type 2 diabetes -related dementia and depression is incompletely understood, it is unclear to what extent metformin or sulfonylurea can prevent or delay new-onset cognitive dysfunction. Therefore, this retrospective study compared the long-term exposure effects of metformin and sulfonylurea, focusing on their outcomes of new diagnosis of dementia, anxiety disorder and depression, and all-cause mortality in patients with type 2 diabetes.

## 2. Methods

### 2.1 Study design and population

This study was approved by the Institutional Review Board of the University of Hong Kong/Hospital Authority Hong Kong West Cluster and the Joint Chinese University of Hong Kong – New Territories East Cluster Clinical Research Ethics Committee. This retrospective population-based cohort was designed to identify the long-term treatment effects of metformin versus sulphonylurea on adverse risks of new-onset dementia and new-onset anxiety disorder and depression, and all-cause mortality with a propensity score matching approach. Patient data used in the study were collected from the Clinical Data Analysis and Reporting System (CDARS), a region-wide database that stores electronic patient health records across local hospitals from the Hong Kong Hospital Authority. Comprehensive medical and clinical data, such as demographics, diagnoses, medications and laboratory tests results can be accessed. The system has been previously used by both our team and other teams in Hong Kong for conducting high-quality epidemiological studies (Chou et al., 2022; Lee et al., 2023a; Zhou et al., 2021). Mortality data was obtained from The Hong Kong Death Registry, an official population-based government registry which hosts registered death records of all Hong Kong citizens. Due to the dependence on ICD-9 coding and death registry records, no adjudication of outcomes was performed. However, the coding and records were set by clinicians and administrative staff, albeit not being directly involved in the study.

### 2.2 Confounding variables

The following potential confounding variables were measured and adjusted at cohort entry: demographic characteristics including baseline age and sex; prior comorbidities including hypertension, coronary heart disease, heart failure, hemiplegia or paraplegia, renal diseases, neurologic diseases, osteoporosis diseases, ophthalmic diseases, liver diseases, ventricular tachycardia/fibrillation, anemia, chronic obstructive pulmonary disease (COPD), ischemic heart disease (IHD), peripheral vascular disease (PVD), gastrointestinal bleeding, malignancy, metastatic solid tumor, and obesity before initial exposure of sulphonylurea and metformin drugs were extracted. CAIDE score, CHA-DS-VASc score and Charlson comorbidity index were calculated. Standard ICD-9 codes to identify prior comorbidities and outcomes of new-onset dementia and new-onset anxiety disorder and depression are provided in **Supplementary Table 1**.

Medications were further processed to extract the drug prescription characteristics of sulphonylurea v.s. metformin, ACEI/ARB, beta blockers, calcium channel blockers, diuretics, stains, antidiabetic drugs, lipid-lowering drugs, nitrates, antihypertensive drugs, statins and fibrates, anticoagulants and antiplatelets. Number of diabetes mellitus and the number of CV drugs were also calculated. Laboratory tests included lymphocyte, neutrophil, platelet, potassium, albumin, sodium, urea, creatinine, total protein, alkaline phosphatase, aspartate transaminase, alanine transaminase, bilirubin, triglyceride, low-density lipoprotein (LDL), LDL calculated, total cholesterol, high-density lipoprotein (HDL), glucose, neutrophil lymphocyte ratio (NLR), fasting blood glucose, and HbA1c.

### 2.3 Primary outcomes

Primary outcomes were new-onset dementia and new-onset anxiety disorder and depression, and secondary outcome was all-cause mortality. Patients were followed up to the end points of new-onset dementia/anxiety disorder and depression, mortality or the end of study (2019/12/31).

### 2.4 Propensity score matching

Propensity score matching approach with 1:1 ratio was used to generate the control cohort of metformin v.s. sulphonylurea drug use based on demographics, CAIDE score, CHA-DS-VASc score, Charlson comorbidity index, past comorbidities, other medications, number of anti-diabetes drugs, number of CV drugs and total cholesterol level. Other matching approaches were also used, including propensity score stratification (Austin, 2011), propensity score matching with inverse probability weighting (Austin and Stuart, 2015), and high-dimensional propensity score approach (Schneeweiss et al., 2009).

### 2.5 Statistical analysis

Univariable Cox regression was used to identify the significant predictors of the primary and secondary outcomes. Hazard ratios (HRs) with corresponding 95% CIs and P-values were reported. Cause-specific hazard models and subdistribution hazard models were further conducted to consider possible competing risks.

Statistical descriptive statistics summarized the baseline and clinical characteristics of the patients with type 2 diabetes. Continuous variables were presented as mean (95% confidence interval [CI] or standard deviation [SD]) and categorical variables were presented as count (%). A standardized mean difference (SMD) of less than 0.2 between the treatment groups post-weighting was considered negligible. All significance tests were two-tailed and considered significant if P-values were 0.05. There was no imputation performed for missing data. No blinding was performed for the predictor as the values were obtained from the electronic health records automatically. Data analyses were performed using RStudio software (Version: 1.1.456) and Python (Version: 3.6).

## 3. Results

### 3.1 Basic characteristics

The cohort included 97,283 patients with type 2 diabetes from 1 January to 31 December 2009. A total of 89,771 patients (46% men, mean age at initial drug use: 67 years old [SD: 12], mean follow up duration of 1,579 days [SD: 650]) remained after excluding 7,512 patients with baseline age less than 18 years old (N = 362), with baseline cancer (N = 811), with both metformin and sulphonylurea use (N = 2132), without use of metformin/sulphonylurea (N = 2,840), with prior diagnosis of dementia and Alzheimer’s/anxiety and depression or medications for cognitive dysfunction (N = 1,367) (**Figure 1**). On the different outcomes, 34,287 patients (Incidence Rate [IR]: 38%) died, 3,401 patients (IR: 4.0%) developed dementia and 1,493 patients (IR: 2.0%) developed anxiety and depression diseases.

**Figure 1.**
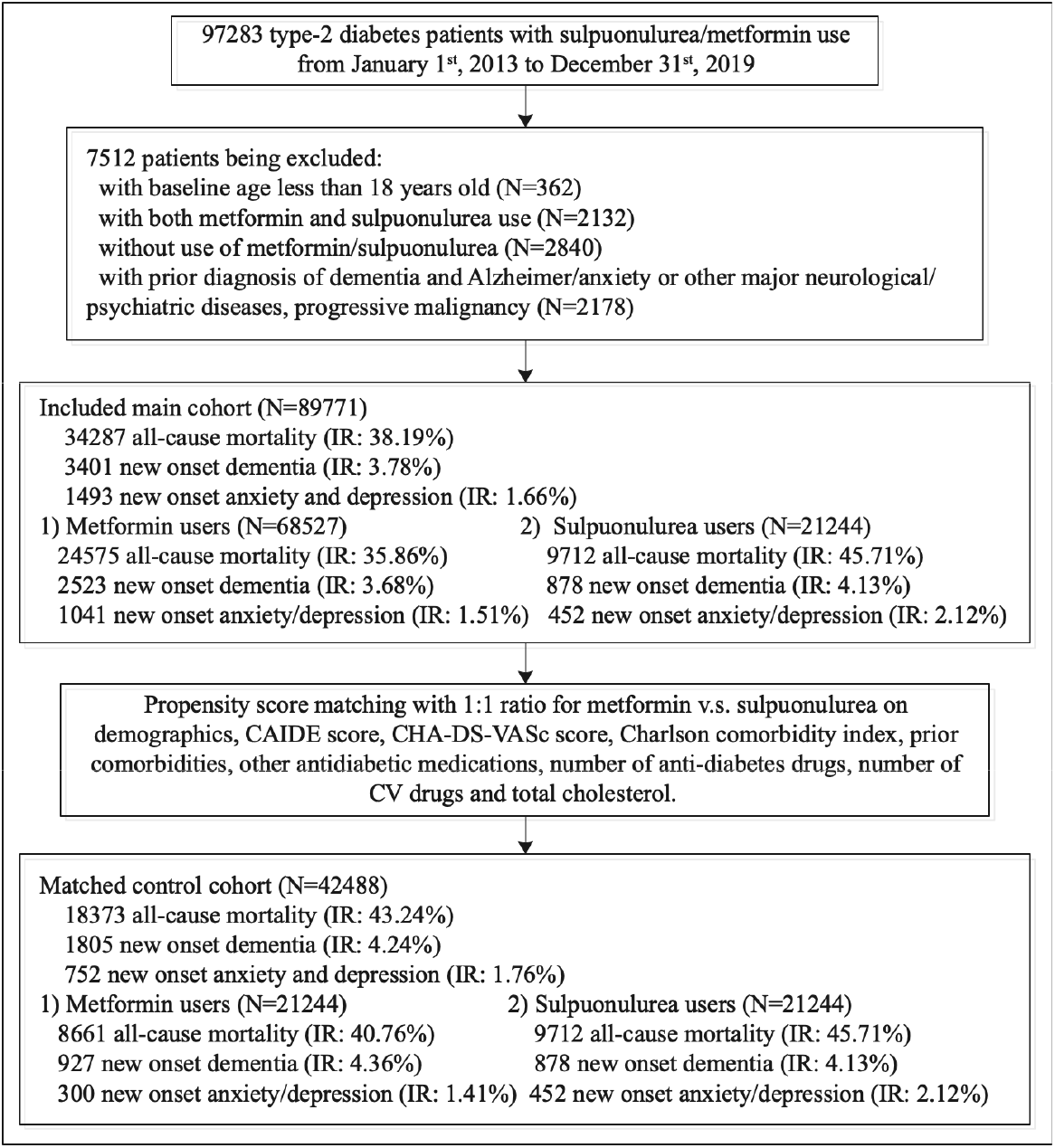
Procedures of constructing study cohorts. IR: incidence rate.

The baseline characteristics between metformin and sulphonylurea users before and after 1:1 propensity score matching are shown in **Table 1**. The technical details of propensity score matching are shown in **Supplementary Tables 2 to 4**. These patients were also categorised into those develop and not developing dementia (**Supplementary Table 5**) and anxiety disorder and depression (**Supplementary Table 6**), and those who died and remained alive at the study-end (**Supplementary Table 7**).

### 3.2 Predictors of primary and secondary outcomes

Univariable Cox regression analysis models were performed to identify the significant risk factors for dementia, anxiety disorder and depression, and all-cause mortality before and after 1:1 propensity score matching as shown in **Table 2** and **Table 3**. Users of metformin showed lower risks of dementia (before: 0.78 [0.72, 0.84], P-value < 0.0001; after: 0.88 [0.80, 0.97], P-value = 0.0074), anxiety disorder and depression (before: 0.77 [0.69, 0.86], P-value < 0.0001; after: 0.71 [0.61, 0.82], P-value < 0.0001), and all-cause mortality (before: 0.69 [0.68, 0.71], P-value < 0.0001; after: 0.83 [0.80, 0.85], P-value < 0.0001). The Kaplan-Meier survival curves of dementia, anxiety disorder and depression, and all-cause mortality for the unmatched and matched cohorts are presented in **Figure 2 and 3**, respectively. The forest plots for new onset dementia and anxiety/depression are shown in **Figure 3**.

**Figure 2.**
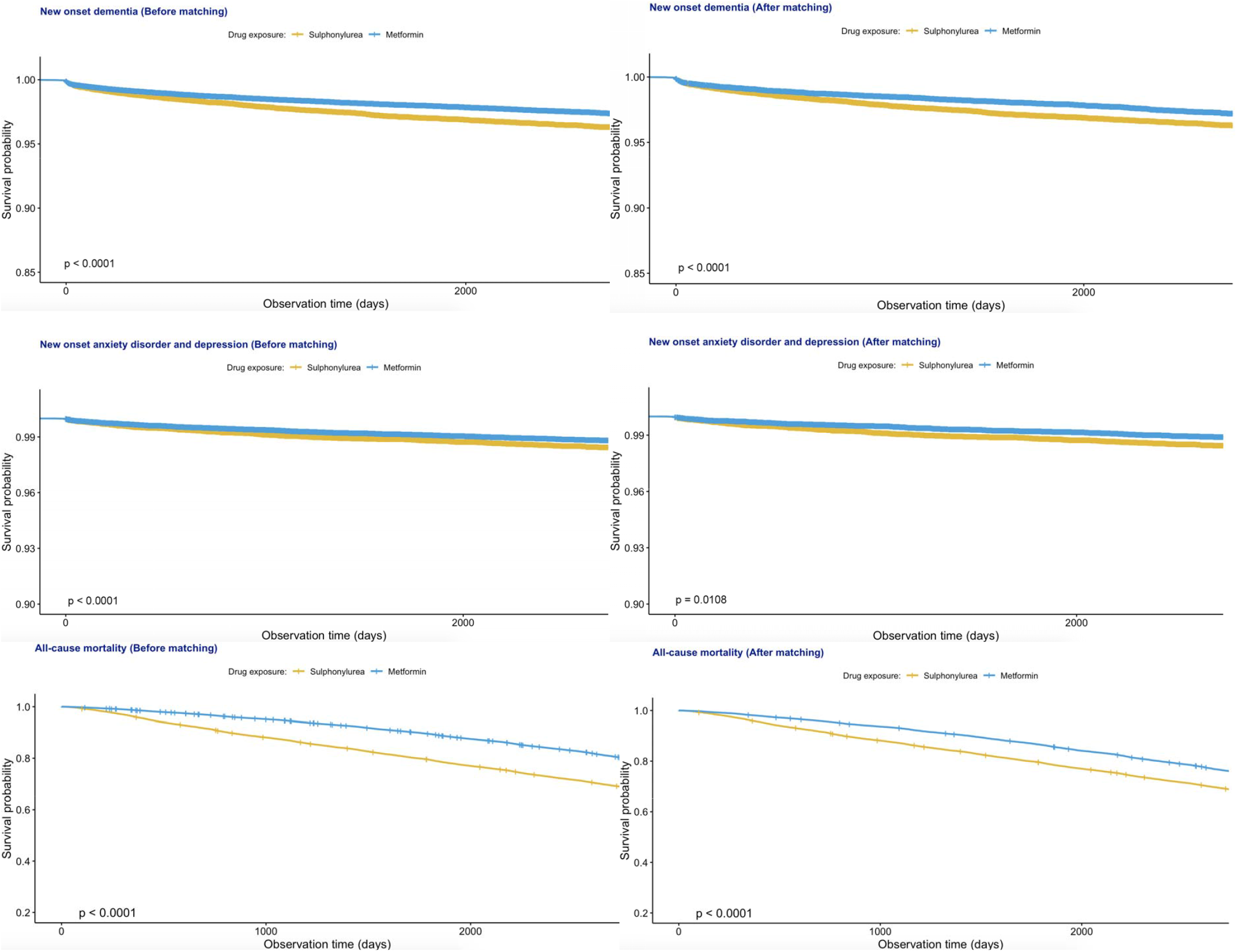
Kaplan Meier survival curves of new onset dementia, anxiety/depression, and all-cause mortality before and after propensity score matching.

**Figure 3.**
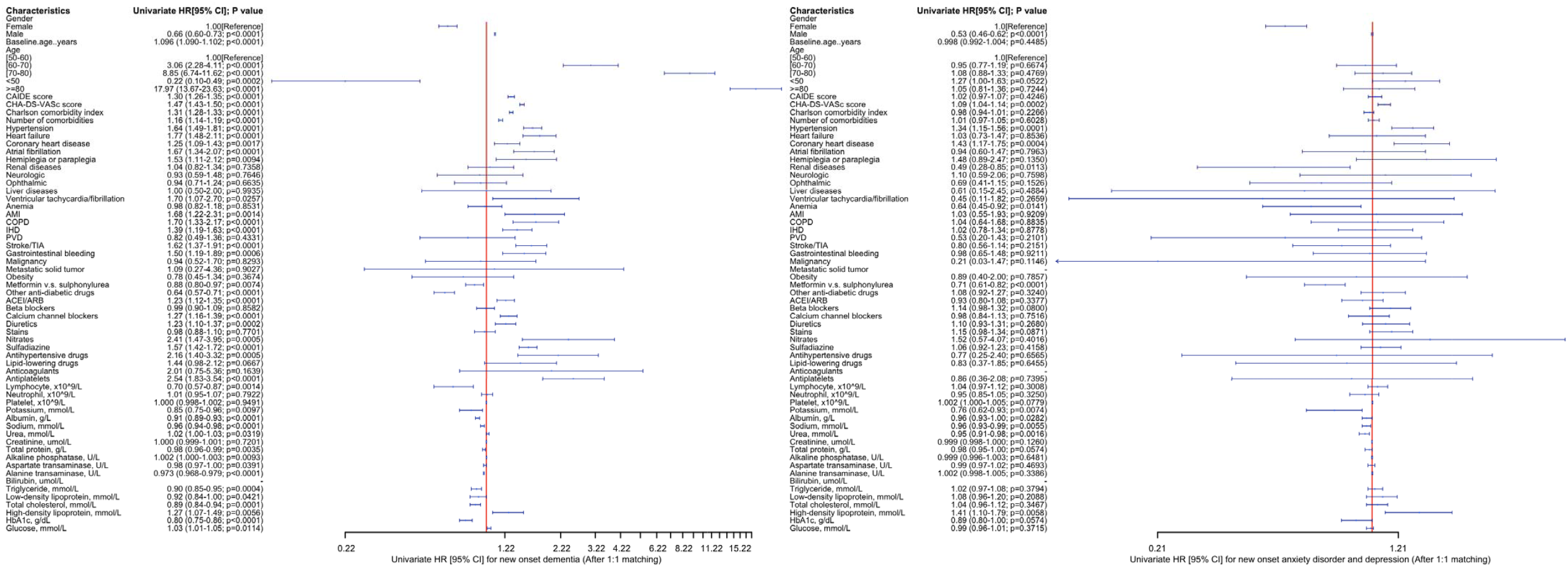
Forest plot portraying the hazard ratios (HR) and 95% CIs of risk factors for new diagnosis of dementia (left), anxiety/depression (right) in the matched cohort.

### 3.3 Competing risk analyses and different approaches using propensity scores

Cause-specific and subdistribution hazard models were applied to confirm the findings from Cox regression, demonstrating that the significant associations between metformin use and the lower risks of the primary and secondary outcomes remained (**Supplementary Table 8**). Risks of new diagnosis of dementia, anxiety disorder/depression and all-cause mortality in matched cohorts associated with for metformin vs. sulphonylurea users with multiple approaches using the propensity score (**Supplementary Table 9**).

## 4. Discussion

This retrospective study compared the use of either metformin or sulphonylurea among patients with type 2 diabetes on the risk of new diagnosis of dementia, anxiety disorder and depression, and all-cause mortality, with preliminary findings presented at a local conference (Hui et al., 2021). We found that: 1) those who developed dementia had lower levels of albumin, alanine transaminase and HbA1c compared to those who developed anxiety disorder and depression; 2) metformin, compared to sulphonylurea, was associated with a lower incidence of dementia, anxiety disorder and depression, and all-cause mortality.

We found that patients with dementia or anxiety disorder and depression had lower levels of albumin, alanine transaminase and HbA1c, which was more pronounced in patients with dementia. Our results are consistent with studies where hypoglycemia was correlated with neurocognitive decline (Sheen and Sheu, 2016). A study previously found that a larger reduction in HbA1c was associated with a greater incidence of dementia (Lee et al., 2019), which was reflected in our univariate Cox regression both before and after propensity score matching. Our findings suggest that appropriate glycemic control and maintenance of normal liver function are important in slowing cognitive decline among patients with diabetes.

Metformin is recommended as the first-line therapy to treat type 2 diabetes in elderly patients (Bosi, 2009). Sulphonylurea, compared to metformin, is associated with an increased risk of hypoglycemia, mortality and cardiovascular events (Maruthur et al., 2016; Whitlock et al., 2020). Metformin has previously shown properties of reducing oxidative stress and restoring memory impairment under diabetic conditions in animal studies (Chen et al., 2016a; Mousavi et al., 2015). These studies agree with our findings, where metformin was associated with a reduced risk of cognitive disorders compared to sulphonylurea.

Several studies have identified anxiety disorder to be more prevalent among patients with type 2 diabetes (Huang et al., 2020; Mezuk et al., 2013; Tu et al., 2017). It is postulated that anxiety disorder is a product of psychological stress from the diagnosis of type 2 diabetes (Pouwer, 2009), or is a complication from extensive medical and exercise regimens necessary for its daily management (Maes et al., 1996). Anxiety disorder may exacerbate type 2 diabetes by increasing the activity of the hypothalamic-pituitary-adrenal axis (HPA axis), inducing the release of hormones such as glucagon, cortisol and epinephrine and in turn increasing blood glucose levels (Bickett and Tapp, 2016). In our study, we found metformin to be superior to sulphonylurea in reducing the risk of anxiety disorder. Another study suggested that the anxiolytic activity of metformin is attributed to its involvement in the AMPK-dependent autophagy regulation pathway after transient forebrain ischemia (Sarkaki et al., 2015).

The occurrence of depression is known to be two to three times higher among patients with diabetes (Badescu et al., 2016). A study found that the prevalence of depression was still higher in patients with undiagnosed type 2 diabetes compared to patients with normal glucose metabolism, suggesting that it is more related to the pathophysiology of type 2 diabetes rather than awareness of the disease (Chen et al., 2016b). Type 2 diabetes and depression have a synergistic effect on micro- and macrovascular complications, incidence of disability and mortality (Black et al., 2003). Early recognition and treatment of type 2 diabetes with metformin may improve clinical outcomes.

### 4.1 Strengths and limitations

To our best acknowledgement, this is the first study to investigate the effects of metformin and sulfonylurea on anxiety disorder and depression. This study has several strengths. We used a large population-based dataset with many clinical and biochemical parameters. The regression was also confirmed using different hazard models, including cause-specific hazard models and subdistribution hazard models. The long follow-up period allows capturing of temporal variability and long-term outcomes.

Several limitations should be noted. This is a retrospective study, thus we cannot analyse clinical outcomes in real-time follow-up. The cohort is recruited from a single region such that external validation is needed to take into account geographical heterogeneity. Furthermore, this retrospective study can only show the association but not causation between metformin and cognitive outcomes.

### 4.2 Conclusion

Metformin use is associated with lower risks of dementia, new-onset anxiety disorder and depression, and all-cause mortality, compared to sulphonylurea use. The protective effects of metformin and possible use in drug repurposing for indications beyond diabetes warrant further investigation.

## Data Availability

All data produced in the present study are available upon reasonable request to the authors

## Declarations

An earlier version of this work has been presented at a local public health conference (Hui JMH et al. AB023. Metformin versus sulfonylurea use for new-onset dementia and anxiety disorder and depression: a propensity score-matched population-based cohort study with competing risk analyses. J Public Health Emerg 2021;5:AB023.), and deposited in the preprint server, MedRxiv.

## Acknowledgements

None.

## Author contributions

JMHH, JZ: data analysis, data interpretation, statistical analysis, manuscript drafting, critical revision of manuscript.

TTLL, KH, OHIC, KSKL, SL: data acquisition and interpretation, critical revision of manuscript.

WTW, AKCW, CC: project planning, data acquisition, data interpretation, critical revision of manuscript.

GT, TL: study conception, study supervision, project planning, data interpretation, statistical analysis, manuscript drafting, critical revision of manuscript.

## References

Austin, P.C., 2011. An Introduction to Propensity Score Methods for Reducing the Effects of Confounding in Observational Studies. Multivariate Behav Res 46, 399–424.

Austin, P.C., Stuart, E.A., 2015. Moving towards best practice when using inverse probability of treatment weighting (IPTW) using the propensity score to estimate causal treatment effects in observational studies. Stat Med 34, 3661–3679.

Badescu, S.V., Tataru, C., Kobylinska, L., Georgescu, E.L., Zahiu, D.M., Zagrean, A.M., Zagrean, L., 2016. The association between Diabetes mellitus and Depression. J Med Life 9, 120–125.

Bickett, A., Tapp, H., 2016. Anxiety and diabetes: Innovative approaches to management in primary care. Exp Biol Med (Maywood) 241, 1724–1731.

Black, S.A., Markides, K.S., Ray, L.A., 2003. Depression predicts increased incidence of adverse health outcomes in older Mexican Americans with type 2 diabetes. Diabetes Care 26, 2822–2828.

Boussageon, R., Bejan-Angoulvant, T., Saadatian-Elahi, M., Lafont, S., Bergeonneau, C., Kassai, B., Erpeldinger, S., Wright, J.M., Gueyffier, F., Cornu, C., 2011. Effect of intensive glucose lowering treatment on all cause mortality, cardiovascular death, and microvascular events in type 2 diabetes: meta-analysis of randomised controlled trials. BMJ 343, d4169.

Chen, F., Dong, R.R., Zhong, K.L., Ghosh, A., Tang, S.S., Long, Y., Hu, M., Miao, M.X., Liao, J.M., Sun, H.B., Kong, L.Y., Hong, H., 2016a. Antidiabetic drugs restore abnormal transport of amyloidbeta across the blood-brain barrier and memory impairment in db/db mice. Neuropharmacology 101, 123–136.

Chen, S., Zhang, Q., Dai, G., Hu, J., Zhu, C., Su, L., Wu, X., 2016b. Association of depression with pre-diabetes, undiagnosed diabetes, and previously diagnosed diabetes: a meta-analysis. Endocrine 53, 35–46.

Cheng, G., Huang, C., Deng, H., Wang, H., 2012. Diabetes as a risk factor for dementia and mild cognitive impairment: a meta-analysis of longitudinal studies. Intern Med J 42, 484–491.

Chou, O.H.I., Zhou, J., Lee, T.T.L., Kot, T., Lee, S., Wai, A.K.C., Wong, W.T., Zhang, Q., Cheng, S.H., Liu, T., Vassiliou, V.S., Cheung, B.M.Y., Tse, G., 2022. Comparisons of the risk of myopericarditis between COVID-19 patients and individuals receiving COVID-19 vaccines: a population-based study. Clin Res Cardiol 111, 1098–1103.

De Silva, T.M., Faraci, F.M., 2016. Microvascular Dysfunction and Cognitive Impairment. Cell Mol Neurobiol 36, 241–258.

Huang, C.J., Hsieh, H.M., Tu, H.P., Jiang, H.J., Wang, P.W., Lin, C.H., 2020. Generalized anxiety disorder in type 2 diabetes mellitus: prevalence and clinical characteristics. Braz J Psychiatry 42, 621–629.

Hui, J.M.H., Zhou, J., Lee, T.T.L., Hui, K., Chou, O.H.I., Lee, Y.H.A., Wai, A.K.C., Jeevaratnam, K., Chang, C., Liu, T., Tse, G., 2021. AB023. Metformin versus sulfonylurea use for new-onset dementia and anxiety disorder and depression: a propensity score-matched population-based cohort study with competing risk analyses. Journal of Public Health and Emergency 5.

Khan, M.A.B., Hashim, M.J., King, J.K., Govender, R.D., Mustafa, H., Al Kaabi, J., 2020. Epidemiology of Type 2 Diabetes - Global Burden of Disease and Forecasted Trends. J Epidemiol Glob Health 10, 107–111.

Lee, A.T.C., Richards, M., Chan, W.C., Chiu, H.F.K., Lee, R.S.Y., Lam, L.C.W., 2019. Higher dementia incidence in older adults with type 2 diabetes and large reduction in HbA1c. Age Ageing 48, 838–844.

Lee, S., Zhou, J., Leung, K.S.K., Wai, A.K.C., Jeevaratnam, K., King, E., Liu, T., Wong, W.T., Chang, C., Wong, I.C.K., Cheung, B.M.Y., Tse, G., Zhang, Q., 2023a. Comparison of Sodium-Glucose Cotransporter-2 Inhibitor and Dipeptidyl Peptidase-4 Inhibitor on the Risks of New-Onset Atrial Fibrillation, Stroke and Mortality in Diabetic Patients: A Propensity Score-Matched Study in Hong Kong. Cardiovasc Drugs Ther 37, 561–569.

Lee, T.T.L., Hui, J.M.H., Lee, Y.H.A., Satti, D.I., Shum, Y.K.L., Kiu, P.T.H., Wai, A.K.C., Liu, T., Wong, W.T., Chan, J.S.K., Cheung, B.M.Y., Wong, I.C.K., Cheng, S.H., Tse, G., 2022a. Sulfonylurea Is Associated With Higher Risks of Ventricular Arrhythmia or Sudden Cardiac Death Compared With Metformin: A Population-Based Cohort Study. J Am Heart Assoc 11, e026289.

Lee, Y.H.A., Hui, J.M.H., Chan, J.S.K., Liu, K., Dee, E.C., Ng, K., Tang, P., Tse, G., Ng, C.F., 2023b. Metformin use and mortality in Asian, diabetic patients with prostate cancer on androgen deprivation therapy: A population-based study. Prostate 83, 119–127.

Lee, Y.H.A., Zhou, J., Hui, J.M.H., Liu, X., Lee, T.T.L., Hui, K., Chan, J.S.K., Wai, A.K.C., Wong, W.T., Liu, T., Ng, K., Lee, S., Dee, E.C., Zhang, Q., Tse, G., 2022b. Risk of New-Onset Prostate Cancer for Metformin Versus Sulfonylurea Use in Type 2 Diabetes Mellitus: A Propensity Score-Matched Study. J Natl Compr Canc Netw 20, 674–682 e615.

MacCallum, L., Senior, P.A., 2019. Safe Use of Metformin in Adults With Type 2 Diabetes and Chronic Kidney Disease: Lower Dosages and Sick-Day Education Are Essential. Can J Diabetes 43, 76–80.

Maes, S., Leventhal, H., de Ridder, D.T.D., 1996. Coping with chronic diseases, Handbook of coping: Theory, research, applications. John Wiley & Sons, Oxford, England, pp. 221–251.

Maruthur, N.M., Tseng, E., Hutfless, S., Wilson, L.M., Suarez-Cuervo, C., Berger, Z., Chu, Y., Iyoha, E., Segal, J.B., Bolen, S., 2016. Diabetes Medications as Monotherapy or Metformin-Based Combination Therapy for Type 2 Diabetes: A Systematic Review and Meta-analysis. Ann Intern Med 164, 740–751.

Mezuk, B., Chen, Y., Yu, C., Guo, Y., Bian, Z., Collins, R., Chen, J., Pang, Z., Wang, H., Peto, R., Que, X., Zhang, H., Tan, Z., Kendler, K.S., Li, L., Chen, Z., 2013. Depression, anxiety, and prevalent diabetes in the Chinese population: findings from the China Kadoorie Biobank of 0.5 million people. J Psychosom Res 75, 511–517.

Mousavi, S.M., Niazmand, S., Hosseini, M., Hassanzadeh, Z., Sadeghnia, H.R., Vafaee, F., Keshavarzi, Z., 2015. Beneficial Effects of Teucrium polium and Metformin on Diabetes-Induced Memory Impairments and Brain Tissue Oxidative Damage in Rats. Int J Alzheimers Dis 2015, 493729.

Pouwer, F., 2009. Should we screen for emotional distress in type 2 diabetes mellitus? Nat Rev Endocrinol 5, 665–671.

Sarkaki, A., Farbood, Y., Badavi, M., Khalaj, L., Khodagholi, F., Ashabi, G., 2015. Metformin improves anxiety-like behaviors through AMPK-dependent regulation of autophagy following transient forebrain ischemia. Metab Brain Dis 30, 1139–1150.

Schneeweiss, S., Rassen, J.A., Glynn, R.J., Avorn, J., Mogun, H., Brookhart, M.A., 2009. Highdimensional propensity score adjustment in studies of treatment effects using health care claims data. Epidemiology 20, 512–522.

Sheen, Y.J., Sheu, W.H., 2016. Association between hypoglycemia and dementia in patients with type 2 diabetes. Diabetes Res Clin Pract 116, 279–287.

Sierra, C., 2012. Cerebral small vessel disease, cognitive impairment and vascular dementia. Panminerva Med 54, 179–188.

Sola, D., Rossi, L., Schianca, G.P., Maffioli, P., Bigliocca, M., Mella, R., Corliano, F., Fra, G.P., Bartoli, E., Derosa, G., 2015. Sulfonylureas and their use in clinical practice. Arch Med Sci 11, 840–848.

Srikanth, V., Sinclair, A.J., Hill-Briggs, F., Moran, C., Biessels, G.J., 2020. Type 2 diabetes and cognitive dysfunction-towards effective management of both comorbidities. Lancet Diabetes Endocrinol 8, 535–545.

Tu, H.P., Lin, C.H., Hsieh, H.M., Jiang, H.J., Wang, P.W., Huang, C.J., 2017. Prevalence of anxiety disorder in patients with type 2 diabetes: a nationwide population-based study in Taiwan 2000-2010. Psychiatr Q 88, 75–91.

van Sloten, T.T., Sedaghat, S., Carnethon, M.R., Launer, L.J., Stehouwer, C.D.A., 2020. Cerebral microvascular complications of type 2 diabetes: stroke, cognitive dysfunction, and depression. Lancet Diabetes Endocrinol 8, 325–336.

Whitlock, R.H., Hougen, I., Komenda, P., Rigatto, C., Clemens, K.K., Tangri, N., 2020. A Safety Comparison of Metformin vs Sulfonylurea Initiation in Patients With Type 2 Diabetes and Chronic Kidney Disease: A Retrospective Cohort Study. Mayo Clin Proc 95, 90–100.

Whitmer, R.A., 2007. Type 2 diabetes and risk of cognitive impairment and dementia. Curr Neurol Neurosci Rep 7, 373–380.

Zhang, Q.Q., Li, W.S., Liu, Z., Zhang, H.L., Ba, Y.G., Zhang, R.X., 2020. Metformin therapy and cognitive dysfunction in patients with type 2 diabetes: A meta-analysis and systematic review. Medicine (Baltimore) 99, e19378.

Zhou, J., Lee, S., Wang, X., Li, Y., Wu, W.K.K., Liu, T., Cao, Z., Zeng, D.D., Leung, K.S.K., Wai, A.K.C., Wong, I.C.K., Cheung, B.M.Y., Zhang, Q., Tse, G., 2021. Development of a multivariable prediction model for severe COVID-19 disease: a population-based study from Hong Kong. NPJ Digit Med 4, 66.

Zhou, J., Zhang, G., Chang, C., Chou, O.H.I., Lee, S., Leung, K.S.K., Wong, W.T., Liu, T., Wai, A.K.C., Cheng, S.H., Zhang, Q., Tse, G., 2022. Metformin versus sulphonylureas for new onset atrial fibrillation and stroke in type 2 diabetes mellitus: a population-based study. Acta Diabetol 59, 697–709.

