## Supplementary material for "Metformin use is associated with lower risks of dementia, anxiety and depression: The Hong Kong Diabetes Study": Table 1

Table 1. Baseline and clinical characteristics of patients with type 2 diabetes mellitus using metformin or sulphonylurea before and after 1:1 propensity score matching.

* for SMD≥0.2

| **Before matching**  **Characteristics Metformin (N=68527)** | **Sulphonylurea (N=21244)** | **SMD** | **After 1:1 matching**  **Metformin (N=21244)** | **Sulphonylurea (N=21244)** | **SMD** |
| --- | --- | --- | --- | --- | --- |
| **Mean(SD);N or Count(%)** | **Mean(SD);N or Count(%)** |  | **Mean(SD);N or Count(%)** | **Mean(SD);N or Count(%)** |  |
| ***Demographics***  Male gender 30413(44.38%) | 10545(49.63%) | 0.11 | 9609(45.23%) | 10545(49.63%) | 0.09 |
| Female gender 38114(55.61%) | 10699(50.36%) | 0.11 | 11635(54.76%) | 10699(50.36%) | 0.09 |
| Baseline age, 66.3(12.1);n=68527 | 66.8(13.7);n=21244 | 0.04 | 67.9(12.0);n=21244 | 66.8(13.7);n=21244 | 0.09 |
| <50 6768(9.87%) | 2598(12.22%) | 0.08 | 1745(8.21%) | 2598(12.22%) | 0.13 |
| [50, 60) 14243(20.78%) | 4031(18.97%) | 0.05 | 3838(18.06%) | 4031(18.97%) | 0.02 |
| [60, 70) 17519(25.56%) | 4598(21.64%) | 0.09 | 5093(23.97%) | 4598(21.64%) | 0.06 |
| [70, 80) 20830(30.39%) | 6094(28.68%) | 0.04 | 7014(33.01%) | 6094(28.68%) | 0.09 |
| >=80 9167(13.37%) | 3923(18.46%) | 0.14 | 3554(16.72%) | 3923(18.46%) | 0.05 |
| ***Past comorbidities*** |  |  |  |  |  |
| CAIDE score 4.3(1.5);n=68527 | 4.3(1.6);n=21244 | 0 | 4.6(1.4);n=21244 | 4.3(1.6);n=21244 | 0.19 |
| CHA-DS-VASc 2.2(1.6);n=68527 | 2.1(1.6);n=21244 | 0.02 | 2.5(1.7);n=21244 | 2.1(1.6);n=21244 | 0.23* |
| Charlson  comorbidity 3.0(2.0);n=68527 | 3.0(2.0);n=21244 | 0.02 | 3.4(2.2);n=21244 | 3.0(2.0);n=21244 | 0.19 |
| index |  |  |  |  |  |
| Number of 1.5(2.0);n=68527 | 1.6(2.1);n=21244 | 0.03 | 2.1(2.2);n=21244 | 1.6(2.1);n=21244 | 0.22* |
| Hypertension 18157(26.49%) | 6477(30.48%) | 0.09 | 8023(37.76%) | 6477(30.48%) | 0.15 |
| Heart failure 3092(4.51%) | 1015(4.77%) | 0.01 | 1524(7.17%) | 1015(4.77%) | 0.1 |
| Coronary heart 5830(8.50%) | 2249(10.58%) | 0.07 | 3104(14.61%) | 2249(10.58%) | 0.12 |
| Atrial 2109(3.07%) | 616(2.89%) | 0.01 | 1019(4.79%) | 616(2.89%) | 0.1 |

years

score

comorbidities

disease fibrillation

| 1053(1.53%) | 289(1.36%) | 0.01 | 503(2.36%) | 289(1.36%) | 0.07 |
| --- | --- | --- | --- | --- | --- |
| 2117(3.08%) | 938(4.41%) | 0.07 | 1084(5.10%) | 938(4.41%) | 0.03 |
| 316(0.46%) | 410(1.92%) | 0.14 | 239(1.12%) | 410(1.92%) | 0.07 |
| 1116(1.62%) | 1071(5.04%) | 0.19 | 767(3.61%) | 1071(5.04%) | 0.07 |
| 246(0.35%) | 94(0.44%) | 0.01 | 151(0.71%) | 94(0.44%) | 0.04 |
| 437(0.63%) | 120(0.56%) | 0.01 | 215(1.01%) | 120(0.56%) | 0.05 |
| 2017(2.94%) | 3037(14.29%) | 0.41* | 1398(6.58%) | 3037(14.29%) | 0.25* |
| 1104(1.61%) | 225(1.05%) | 0.05 | 442(2.08%) | 225(1.05%) | 0.08 |
| 1609(2.34%) | 431(2.02%) | 0.02 | 755(3.55%) | 431(2.02%) | 0.09 |
| 5487(8.00%) | 1540(7.24%) | 0.03 | 2459(11.57%) | 1540(7.24%) | 0.15 |
| 681(0.99%) | 233(1.09%) | 0.01 | 366(1.72%) | 233(1.09%) | 0.05 |
| 4120(6.01%) | 1116(5.25%) | 0.03 | 1844(8.68%) | 1116(5.25%) | 0.13 |
| 1895(2.76%) | 580(2.73%) | 0 | 976(4.59%) | 580(2.73%) | 0.1 |
| 438(0.63%) | 112(0.52%) | 0.01 | 204(0.96%) | 112(0.52%) | 0.05 |
| 103(0.15%) | 18(0.08%) | 0.02 | 43(0.20%) | 18(0.08%) | 0.03 |
| 937(1.36%) | 146(0.68%) | 0.07 | 284(1.33%) | 146(0.68%) | 0.06 |
| 18163(26.50%) | 6957(32.74%) | 0.14 | 5304(24.96%) | 6957(32.74%) | 0.17 |
| 36691(53.54%) | 7576(35.66%) | 0.37* | 11881(55.92%) | 7576(35.66%) | 0.42* |
| 24324(35.49%) | 6718(31.62%) | 0.08 | 8959(42.17%) | 6718(31.62%) | 0.22* |
| 30405(44.36%) | 8109(38.17%) | 0.13 | 10309(48.52%) | 8109(38.17%) | 0.21* |
| 12744(18.59%) | 4655(21.91%) | 0.08 | 5331(25.09%) | 4655(21.91%) | 0.08 |
| 17819(26.00%) | 4541(21.37%) | 0.11 | 6749(31.76%) | 4541(21.37%) | 0.24* |
| 135(0.19%) | 148(0.69%) | 0.07 | 108(0.50%) | 148(0.69%) | 0.02 |
| 51612(75.31%) | 9027(42.49%) | 0.71* | 12485(58.76%) | 9027(42.49%) | 0.33* |

Hemiplegia or paraplegia Renal diseases Neurologic Ophthalmic Liver diseases Ventricular tachycardia/fibr illation

Anemia AMI COPD IHD PVD

Stroke/TIA Gastrointestinal bleeding Malignancy Metastatic solid tumor

Obesity ***Medications*** Other anti- diabetic drugs ACEI/ARB

Beta blockers Calcium channel blockers Diuretics Stains Nitrates Sulfadiazine

| 168(0.24%) | 200(0.94%) | 0.09 | 140(0.65%) | 200(0.94%) | 0.03 |
| --- | --- | --- | --- | --- | --- |
| 367(0.53%) | 318(1.49%) | 0.1 | 270(1.27%) | 318(1.49%) | 0.02 |
| 26(0.03%) | 53(0.24%) | 0.06 | 26(0.12%) | 53(0.24%) | 0.03 |
| 293(0.42%) | 291(1.36%) | 0.1 | 221(1.04%) | 291(1.36%) | 0.03 |
| 2.0(1.0);n=4819 | 1.7(1.7);n=5126 | 0.21* | 1.9(0.7);n=2185 | 1.7(1.7);n=5126 | 0.15 |
| 5.3(2.5);n=4819 | 5.6(2.9);n=5126 | 0.12 | 5.5(2.5);n=2185 | 5.6(2.9);n=5126 | 0.06 |
| 260.2(84.4);n=5074 | 253.9(88.4);n=648 | 0.07 | 257.2(80.8);n=1844 | 253.9(88.4);n=648 | 0.04 |
| 4.3(0.5);n=40865 | 4.2(0.5);n=9536 | 0.07 | 4.2(0.5);n=13723 | 4.2(0.5);n=9536 | 0.01 |
| 39.3(4.7);n=10335 | 37.6(5.8);n=6269 | 0.32* | 38.8(4.9);n=3942 | 37.6(5.8);n=6269 | 0.22* |
| 139.3(3.1);n=40883 | 139.2(3.4);n=9545 | 0.04 | 139.4(3.2);n=13732 | 139.2(3.4);n=9545 | 0.07 |
| 6.2(2.4);n=35998 | 9.4(6.8);n=9032 | 0.63* | 6.4(2.7);n=12201 | 9.4(6.8);n=9032 | 0.57* |
| 87.3(30.1);n=36201 | 166.4(196.5);n=9076 | 0.56* | 91.5(34.7);n=12280 | 166.4(196.5);n=9076 | 0.53* |
| 74.7(6.3);n=10309 | 73.9(7.5);n=6243 | 0.12 | 74.4(6.6);n=3930 | 73.9(7.5);n=6243 | 0.08 |
| 75.2(31.1);n=33237 | 88.7(46.0);n=8713 | 0.34* | 76.3(30.1);n=11349 | 88.7(46.0);n=8713 | 0.32* |
| 30.7(72.4);n=2380 | 34.9(61.2);n=927 | 0.06 | 31.5(66.3);n=784 | 34.9(61.2);n=927 | 0.05 |
| 24.8(22.0);n=26271 | 26.3(26.8);n=7224 | 0.06 | 24.9(19.8);n=9317 | 26.3(26.8);n=7224 | 0.06 |
| 10.5(4.3);n=10 | 13.4(11.6);n=1980 | 0.33* | 10.2(4.3);n=10 | 13.4(11.6);n=1980 | 0.37* |

Antihypertensiv e drugs

Lipid-lowering drugs Anticoagulants Antiplatelets ***Laboratory tests*** Lymphocyte, x10^9/L Neutrophil,

x10^9/L Platelet, x10^9/L Potassium, mmol/L Albumin, g/L Sodium, mmol/L

Urea, mmol/L Creatinine, umol/L

Total protein,

g/L Alkaline

phosphatase, U/L Aspartate transaminase, U/L

Alanine transaminase, U/L Bilirubin, umol/L

mmol/L

| Triglyceride, 1.7(1.3);n=50770 | 1.7(1.6);n=14286 | 0.01 | 1.8(1.2);n=16544 | 1.7(1.6);n=14286 | 0.03 |
| --- | --- | --- | --- | --- | --- |
| Low-density  lipoprotein, 2.9(0.8);n=32742 | 3.0(0.9);n=8350 | 0.07 | 2.9(0.9);n=10850 | 3.0(0.9);n=8350 | 0.07 |
| mmol/L  Total  cholesterol, 4.8(1.0);n=50858 | 4.9(1.1);n=14348 | 0.12 | 4.8(1.0);n=16569 | 4.9(1.1);n=14348 | 0.13 |
| mmol/L  High-density  lipoprotein, 1.2(0.3);n=46097 | 1.2(0.4);n=11552 | 0.1 | 1.2(0.3);n=15025 | 1.2(0.4);n=11552 | 0.11 |
| mmol/L |  |  |  |  |  |
| HbA1c, g/dL 12.8(1.7);n=5526 | 12.1(2.2);n=5711 | 0.37* | 12.3(1.7);n=2437 | 12.1(2.2);n=5711 | 0.07 |
| Glucose, 9.1(4.1);n=18817 | 8.4(4.3);n=10522 | 0.17 | 8.5(3.8);n=10329 | 8.4(4.3);n=10522 | 0.02 |

mmol/L

CAIDE: cardiovascular risk factors, aging, and incidence of dementia, AMI: acute myocardial infarction, COPD: chronic obstructive pulmonary disease, IHD: ischemic heart disease, PVD: peripheral vascular disease, TIA: transient ischemic attack, ACEI: angiotensin-converting-enzyme inhibitors, ARB: angiotensin II receptor blockers, LDL: low density lipoprotein cholesterol, HDL: high density lipoprotein cholesterol, SD: standard deviation, SMD: standard mean difference.
