## Supplementary material for "Metformin use is associated with lower risks of dementia, anxiety and depression: The Hong Kong Diabetes Study": Table 2

Table 2. Univariable Cox regression for new diagnosis of dementia or anxiety/depression before and after 1:1 propensity score matching.

|  | **Before matching** |  | **After 1:1 matching** |  |
| --- | --- | --- | --- | --- |
| **Characteristics** | **Dementia**  **HR [95% CI]; P value** | **Anxiety disorder and depression HR [95% CI]; P value** | **Dementia**  **HR [95% CI]; P value** | **Anxiety disorder and depression [95% CI]; P value** |
| ***Demographics*** |  |  |  |  |
| Gender |  |  |  |  |
| Female | - | - | - | - |
| Male | 0.63 (0.58-0.67; p<0.0001) | 0.54 (0.48-0.60; p<0.0001) | 0.66 (0.60-0.73; p<0.0001) | 0.53 (0.46-0.62; p<0.0001) |
| Baseline age, years | 1.098 (1.094-1.102; p<0.0001) | 0.995 (0.991-1.000; p=0.0310) | 1.096 (1.090-1.102; p<0.0001) | 0.998 (0.992-1.004; p=0.4485) |
| Age |  |  |  |  |
| [50-60) | - | - | - | - |
| [60-70) | 4.07 (3.26-5.07; p<0.0001) | 0.90 (0.77-1.04; p=0.1453) | 3.06 (2.28-4.11; p<0.0001) | 0.95 (0.77-1.19; p=0.6674) |
| [70-80) | 11.42 (9.28-14.06; p<0.0001) | 0.92 (0.79-1.06; p=0.2342) | 8.85 (6.74-11.62; p<0.0001) | 1.08 (0.88-1.33; p=0.4769) |
| <50 | 0.20 (0.11-0.39; p<0.0001) | 1.07 (0.90-1.27; p=0.4663) | 0.22 (0.10-0.49; p=0.0002) | 1.27 (1.00-1.63; p=0.0522) |
| >=80 | 22.24 (18.03-27.43; p<0.0001) | 0.94 (0.78-1.13; p=0.5075) | 17.97 (13.67-23.63; p<0.0001) | 1.05 (0.81-1.36; p=0.7244) |
| ***Past***  ***comorbidities*** |  |  |  |  |
| CAIDE score | 1.32 (1.29-1.36; p<0.0001) | 1.03 (1.00-1.07; p=0.0593) | 1.30 (1.26-1.35; p<0.0001) | 1.02 (0.97-1.07; p=0.4246) |
| CHA-DS-VASc  score | 1.48 (1.45-1.51; p<0.0001) | 1.09 (1.05-1.12; p<0.0001) | 1.47 (1.43-1.50; p<0.0001) | 1.09 (1.04-1.14; p=0.0002) |
| Charlson  comorbidity index | 1.31 (1.30-1.33; p<0.0001) | 1.00 (0.97-1.02; p=0.7198) | 1.31 (1.28-1.33; p<0.0001) | 0.98 (0.94-1.01; p=0.2266) |
| Number of  comorbidities | 1.15 (1.14-1.17; p<0.0001) | 1.04 (1.01-1.07; p=0.0055) | 1.16 (1.14-1.19; p<0.0001) | 1.01 (0.97-1.05; p=0.6028) |
| Hypertension | 1.67 (1.56-1.79; p<0.0001) | 1.31 (1.17-1.46; p<0.0001) | 1.64 (1.49-1.81; p<0.0001) | 1.34 (1.15-1.56; p=0.0001) |
| Heart failure | 1.68 (1.45-1.95; p<0.0001) | 1.10 (0.84-1.44; p=0.4722) | 1.77 (1.48-2.11; p<0.0001) | 1.03 (0.73-1.47; p=0.8536) |
| Coronary heart disease | 1.29 (1.15-1.45; p<0.0001) | 1.42 (1.21-1.67; p<0.0001) | 1.25 (1.09-1.43; p=0.0017) | 1.43 (1.17-1.75; p=0.0004) |

| Atrial  fibrillation | 1.61 (1.35-1.93; p<0.0001) | 1.17 (0.85-1.60; p=0.3278) | 1.67 (1.34-2.07; p<0.0001) | 0.94 (0.60-1.47; p=0.7963) |
| --- | --- | --- | --- | --- |
| Hemiplegia or paraplegia | 1.44 (1.10-1.87; p=0.0080) | 1.29 (0.85-1.97; p=0.2320) | 1.53 (1.11-2.12; p=0.0094) | 1.48 (0.89-2.47; p=0.1350) |
| Renal diseases | 1.16 (0.95-1.42; p=0.1383) | 0.54 (0.35-0.83; p=0.0049) | 1.04 (0.82-1.34; p=0.7358) | 0.49 (0.28-0.85; p=0.0113) |
| Neurologic | 0.92 (0.58-1.46; p=0.7254) | 1.09 (0.60-1.98; p=0.7726) | 0.93 (0.59-1.48; p=0.7646) | 1.10 (0.59-2.06; p=0.7598) |
| Ophthalmic | 0.92 (0.70-1.21; p=0.5525) | 0.63 (0.38-1.02; p=0.0621) | 0.94 (0.71-1.24; p=0.6635) | 0.69 (0.41-1.15; p=0.1526) |
| Liver diseases | 1.02 (0.55-1.91; p=0.9379) | 0.46 (0.12-1.85; p=0.2761) | 1.00 (0.50-2.00; p=0.9935) | 0.61 (0.15-2.45; p=0.4884) |
| Ventricular  tachycardia/fibri llation | 1.66 (1.13-2.44; p=0.0105) | 0.86 (0.38-1.91; p=0.7022) | 1.70 (1.07-2.70; p=0.0257) | 0.45 (0.11-1.82; p=0.2659) |
| Anemia | 1.06 (0.89-1.26; p=0.5212) | 0.76 (0.56-1.04; p=0.0831) | 0.98 (0.82-1.18; p=0.8531) | 0.64 (0.45-0.92; p=0.0141) |
| AMI | 1.50 (1.16-1.93; p=0.0017) | 0.87 (0.53-1.43; p=0.5822) | 1.68 (1.22-2.31; p=0.0014) | 1.03 (0.55-1.93; p=0.9209) |
| COPD | 1.60 (1.31-1.97; p<0.0001) | 1.34 (0.96-1.87; p=0.0812) | 1.70 (1.33-2.17; p<0.0001) | 1.04 (0.64-1.68; p=0.8835) |
| IHD | 1.33 (1.17-1.50; p<0.0001) | 1.14 (0.94-1.39; p=0.1869) | 1.39 (1.19-1.63; p<0.0001) | 1.02 (0.78-1.34; p=0.8778) |
| PVD | 1.08 (0.75-1.56; p=0.6657) | 0.81 (0.42-1.56; p=0.5330) | 0.82 (0.49-1.36; p=0.4331) | 0.53 (0.20-1.43; p=0.2101) |
| Stroke/TIA | 1.58 (1.38-1.80; p<0.0001) | 0.94 (0.74-1.21; p=0.6503) | 1.62 (1.37-1.91; p<0.0001) | 0.80 (0.56-1.14; p=0.2151) |
| Gastrointestinal bleeding | 1.41 (1.15-1.71; p=0.0007) | 0.90 (0.63-1.27; p=0.5328) | 1.50 (1.19-1.89; p=0.0006) | 0.98 (0.65-1.48; p=0.9211) |
| Malignancy | 0.92 (0.56-1.50; p=0.7406) | 0.13 (0.02-0.91; p=0.0401) | 0.94 (0.52-1.70; p=0.8293) | 0.21 (0.03-1.47; p=0.1146) |
| Metastatic solid tumor | 0.62 (0.16-2.48; p=0.5004) | 0.00 (0.00-121.00; p=0.9717) | 1.09 (0.27-4.36; p=0.9027) | 0.00 (0.00-Inf; p=0.9865) |
| Obesity | 0.52 (0.33-0.81; p=0.0042) | 0.88 (0.52-1.48; p=0.6198) | 0.78 (0.45-1.34; p=0.3674) | 0.89 (0.40-2.00; p=0.7857) |
| ***Medications*** |  |  |  |  |
| Metformin v.s. sulphonylurea | 0.78 (0.72-0.84; p<0.0001) | 0.77 (0.69-0.86; p<0.0001) | 0.88 (0.80-0.97; p=0.0074) | 0.71 (0.61-0.82; p<0.0001) |
| Other anti-  diabetic drugs | 0.66 (0.61-0.72; p<0.0001) | 0.89 (0.79-1.00; p=0.0478) | 0.64 (0.57-0.71; p<0.0001) | 1.08 (0.92-1.27; p=0.3240) |
| ACEI/ARB | 1.06 (0.99-1.14; p=0.0844) | 0.94 (0.85-1.05; p=0.2751) | 1.23 (1.12-1.35; p<0.0001) | 0.93 (0.80-1.08; p=0.3377) |
| Beta blockers | 1.04 (0.97-1.12; p=0.2318) | 1.17 (1.05-1.30; p=0.0036) | 0.99 (0.90-1.09; p=0.8582) | 1.14 (0.98-1.32; p=0.0800) |
| Calcium channel blockers | 1.34 (1.25-1.44; p<0.0001) | 0.93 (0.83-1.03; p=0.1534) | 1.27 (1.16-1.39; p<0.0001) | 0.98 (0.84-1.13; p=0.7516) |
| Diuretics | 1.37 (1.26-1.48; p<0.0001) | 1.08 (0.95-1.24; p=0.2288) | 1.23 (1.10-1.37; p=0.0002) | 1.10 (0.93-1.31; p=0.2680) |

| Stains | 0.88 (0.81-0.95; p=0.0017) | 1.18 (1.05-1.32; p=0.0041) | 0.98 (0.88-1.10; p=0.7701) | 1.15 (0.98-1.34; p=0.0871) |
| --- | --- | --- | --- | --- |
| Nitrates | 2.58 (1.60-4.16; p=0.0001) | 1.76 (0.73-4.24; p=0.2064) | 2.41 (1.47-3.95; p=0.0005) | 1.52 (0.57-4.07; p=0.4016) |
| Sulfadiazine | 1.25 (1.16-1.35; p<0.0001) | 0.97 (0.87-1.08; p=0.5487) | 1.57 (1.42-1.72; p<0.0001) | 1.06 (0.92-1.23; p=0.4158) |
| Antihypertensiv e drugs | 2.24 (1.46-3.44; p=0.0002) | 0.74 (0.24-2.30; p=0.6053) | 2.16 (1.40-3.32; p=0.0005) | 0.77 (0.25-2.40; p=0.6565) |
| Lipid-lowering drugs | 1.44 (0.98-2.10; p=0.0617) | 1.23 (0.66-2.29; p=0.5151) | 1.44 (0.98-2.12; p=0.0667) | 0.83 (0.37-1.85; p=0.6455) |
| Anticoagulants | 2.28 (0.86-6.09; p=0.0991) | 0.00 (0.00-Inf; p=0.9791) | 2.01 (0.75-5.36; p=0.1639) | 0.00 (0.00-Inf; p=0.9859) |
| Antiplatelets | 2.71 (1.97-3.71; p<0.0001) | 1.26 (0.63-2.52; p=0.5165) | 2.54 (1.83-3.54; p<0.0001) | 0.86 (0.36-2.08; p=0.7395) |
| ***Laboratory tests*** |  |  |  |  |
| Lymphocyte, x10^9/L | 0.64 (0.53-0.78; p<0.0001) | 1.05 (1.00-1.10; p=0.0701) | 0.70 (0.57-0.87; p=0.0014) | 1.04 (0.97-1.12; p=0.3008) |
| Neutrophil,  x10^9/L | 1.04 (0.99-1.09; p=0.0875) | 0.94 (0.86-1.02; p=0.1412) | 1.01 (0.95-1.07; p=0.7922) | 0.95 (0.85-1.05; p=0.3250) |
| Platelet,  x10^9/L | 0.999 (0.998-1.000;  p=0.1158) | 1.001 (1.000-1.003; p=0.1173) | 1.000 (0.998-1.002;  p=0.9491) | 1.002 (1.000-1.005; p=0.0779) |
| Potassium, mmol/L | 0.77 (0.70-0.84; p<0.0001) | 0.70 (0.60-0.81; p<0.0001) | 0.85 (0.75-0.96; p=0.0097) | 0.76 (0.62-0.93; p=0.0074) |
| Albumin, g/L | 0.91 (0.89-0.92; p<0.0001) | 0.97 (0.94-0.99; p=0.0074) | 0.91 (0.89-0.93; p<0.0001) | 0.96 (0.93-1.00; p=0.0282) |
| Sodium, mmol/L | 0.99 (0.97-1.00; p=0.0830) | 0.96 (0.94-0.98; p=0.0003) | 0.96 (0.94-0.98; p<0.0001) | 0.96 (0.93-0.99; p=0.0055) |
| Urea, mmol/L | 1.04 (1.03-1.05; p<0.0001) | 0.96 (0.93-0.98; p=0.0017) | 1.02 (1.00-1.03; p=0.0319) | 0.95 (0.91-0.98; p=0.0016) |
| Creatinine, umol/L | 1.001 (1.000-1.001; p=0.0522) | 0.999 (0.997-1.000; p=0.1006) | 1.000 (0.999-1.001; p=0.7201) | 0.999 (0.998-1.000; p=0.1260) |
| Total protein,  g/L | 0.97 (0.96-0.99; p=0.0003) | 0.98 (0.96-1.00; p=0.0928) | 0.98 (0.96-0.99; p=0.0035) | 0.98 (0.95-1.00; p=0.0574) |
| Alkaline  phosphatase, U/L | 1.002 (1.001-1.003; p<0.0001) | 1.001 (0.999-1.003; p=0.3877) | 1.002 (1.000-1.003; p=0.0093) | 0.999 (0.996-1.003; p=0.6481) |
| Aspartate  transaminase, U/L | 1.001 (0.999-1.002; p=0.3926) | 0.99 (0.97-1.01; p=0.1934) | 0.98 (0.97-1.00; p=0.0391) | 0.99 (0.97-1.02; p=0.4693) |

| Alanine transaminase,  U/L | 0.98 (0.97-0.98; p<0.0001) | 1.002 (0.999-1.004; p=0.1463) | 0.973 (0.968-0.979; p<0.0001) | 1.002 (0.998-1.005; p=0.3386) |
| --- | --- | --- | --- | --- |
| Bilirubin,  umol/L | - | NA (NA-NA; p=NA) | - | NA (NA-NA; p=NA) |
| Triglyceride, mmol/L | 0.91 (0.87-0.94; p<0.0001) | 1.06 (1.03-1.08; p<0.0001) | 0.90 (0.85-0.95; p=0.0004) | 1.02 (0.97-1.08; p=0.3794) |
| Low-density lipoprotein,  mmol/L | 0.92 (0.86-0.99; p=0.0165) | 1.09 (1.00-1.18; p=0.0456) | 0.92 (0.84-1.00; p=0.0421) | 1.08 (0.96-1.20; p=0.2088) |
| Total cholesterol,  mmol/L | 0.92 (0.88-0.96; p=0.0001) | 1.09 (1.03-1.15; p=0.0017) | 0.89 (0.84-0.94; p=0.0001) | 1.04 (0.96-1.12; p=0.3467) |
| High-density lipoprotein,  mmol/L | 1.27 (1.12-1.44; p=0.0002) | 1.39 (1.16-1.66; p=0.0003) | 1.27 (1.07-1.49; p=0.0056) | 1.41 (1.10-1.79; p=0.0058) |
| HbA1c, g/dL | 0.78 (0.73-0.83; p<0.0001) | 0.92 (0.83-1.01; p=0.0694) | 0.80 (0.75-0.86; p<0.0001) | 0.89 (0.80-1.00; p=0.0574) |
| Glucose, mmol/L | 1.02 (1.01-1.04; p=0.0072) | 0.98 (0.96-1.00; p=0.0772) | 1.03 (1.01-1.05; p=0.0114) | 0.99 (0.96-1.01; p=0.3715) |

CAIDE: cardiovascular risk factors, aging, and incidence of dementia, AMI: acute myocardial infarction, COPD: chronic obstructive pulmonary disease, IHD: ischemic heart disease, PVD: peripheral vascular disease, TIA: transient ischemic attack, ACEI: angiotensin-converting-enzyme inhibitors, ARB: angiotensin II receptor blockers, LDL: low density lipoprotein cholesterol, HDL: high density lipoprotein cholesterol, HR: hazard ratio, CI: confidence interval.
