## Supplementary material for "Metformin use is associated with lower risks of dementia, anxiety and depression: The Hong Kong Diabetes Study": Table 3

Table 3. Univariable Cox regression for all-cause mortality before and after 1:1 propensity score matching. Characteristics	Before matching	After 1:1 matching

|  | **HR [95% CI]; P value** | **HR [95% CI]; P value** |
| --- | --- | --- |
| ***Demographics*** |  |  |
| Gender |  |  |
| Female | - | - |
| Male | 1.11 (1.08-1.13; p<0.0001) | 1.12 (1.08-1.15; p<0.0001) |
| Baseline age, years | 1.081 (1.080-1.082; p<0.0001) | 1.077 (1.075-1.078; p<0.0001) |
| Age |  |  |
| [50-60) | - | - |
| [60-70) | 1.93 (1.84-2.01; p<0.0001) | 1.79 (1.68-1.90; p<0.0001) |
| [70-80) | 4.29 (4.12-4.46; p<0.0001) | 3.89 (3.68-4.12; p<0.0001) |
| <50 | 0.69 (0.64-0.74; p<0.0001) | 0.62 (0.56-0.69; p<0.0001) |
| >=80 | 9.50 (9.11-9.90; p<0.0001) | 8.24 (7.78-8.73; p<0.0001) |
| ***Past comorbidities*** |  |  |
| CAIDE score | 1.36 (1.35-1.37; p<0.0001) | 1.34 (1.33-1.36; p<0.0001) |
| CHA-DS-VASc score | 1.43 (1.42-1.44; p<0.0001) | 1.40 (1.39-1.41; p<0.0001) |
| Charlson comorbidity index | 1.33 (1.32-1.33; p<0.0001) | 1.305 (1.298-1.313; p<0.0001) |
| Number of comorbidities | 1.21 (1.21-1.22; p<0.0001) | 1.218 (1.211-1.225; p<0.0001) |
| Hypertension | 2.08 (2.04-2.13; p<0.0001) | 2.05 (1.99-2.11; p<0.0001) |
| Heart failure | 2.47 (2.38-2.57; p<0.0001) | 2.31 (2.20-2.42; p<0.0001) |
| Coronary heart disease | 1.77 (1.72-1.83; p<0.0001) | 1.72 (1.66-1.79; p<0.0001) |
| Atrial fibrillation | 2.23 (2.13-2.34; p<0.0001) | 2.04 (1.92-2.16; p<0.0001) |
| Hemiplegia or paraplegia | 2.03 (1.90-2.17; p<0.0001) | 1.93 (1.77-2.10; p<0.0001) |
| Renal diseases | 2.44 (2.34-2.55; p<0.0001) | 2.22 (2.10-2.34; p<0.0001) |
| Neurologic | 2.97 (2.72-3.23; p<0.0001) | 2.69 (2.46-2.94; p<0.0001) |
| Ophthalmic | 2.47 (2.35-2.61; p<0.0001) | 2.36 (2.23-2.49; p<0.0001) |
| Liver diseases | 2.29 (2.01-2.59; p<0.0001) | 1.84 (1.58-2.14; p<0.0001) |
| Ventricular tachycardia/fibrillation | 2.20 (1.99-2.44; p<0.0001) | 1.93 (1.69-2.19; p<0.0001) |

| Anemia | 3.34 (3.23-3.45; p<0.0001) | 3.21 (3.10-3.34; p<0.0001) |
| --- | --- | --- |
| AMI | 1.63 (1.52-1.75; p<0.0001) | 1.39 (1.26-1.54; p<0.0001) |
| COPD | 1.89 (1.79-2.00; p<0.0001) | 1.73 (1.61-1.86; p<0.0001) |
| IHD | 1.67 (1.61-1.72; p<0.0001) | 1.64 (1.57-1.71; p<0.0001) |
| PVD | 2.48 (2.30-2.68; p<0.0001) | 2.16 (1.96-2.37; p<0.0001) |
| Stroke/TIA | 1.82 (1.75-1.89; p<0.0001) | 1.72 (1.63-1.80; p<0.0001) |
| Gastrointestinal bleeding | 1.77 (1.68-1.86; p<0.0001) | 1.61 (1.51-1.72; p<0.0001) |
| Malignancy | 1.27 (1.13-1.44; p=0.0001) | 0.93 (0.78-1.10; p=0.3869) |
| Metastatic solid tumor | 3.12 (2.57-3.80; p<0.0001) | 2.48 (1.87-3.28; p<0.0001) |
| Obesity | 0.86 (0.77-0.95; p=0.0026) | 0.78 (0.66-0.91; p=0.0013) |
| ***Medications*** |  |  |
| Metformin v.s. sulphonylurea | 0.69 (0.68-0.71; p<0.0001) | 0.83 (0.80-0.85; p<0.0001) |
| Other anti-diabetic drugs | 0.94 (0.92-0.96; p<0.0001) | 1.12 (1.08-1.15; p<0.0001) |
| ACEI/ARB | 1.32 (1.29-1.35; p<0.0001) | 1.42 (1.38-1.47; p<0.0001) |
| Beta blockers | 1.26 (1.23-1.29; p<0.0001) | 1.18 (1.15-1.22; p<0.0001) |
| Calcium channel blockers | 1.76 (1.72-1.80; p<0.0001) | 1.68 (1.63-1.73; p<0.0001) |
| Diuretics | 1.79 (1.75-1.83; p<0.0001) | 1.76 (1.71-1.82; p<0.0001) |
| Stains | 1.16 (1.13-1.19; p<0.0001) | 1.20 (1.16-1.24; p<0.0001) |
| Nitrates | 3.47 (3.03-3.97; p<0.0001) | 3.00 (2.61-3.46; p<0.0001) |
| Sulfadiazine | 1.03 (1.01-1.05; p=0.0112) | 1.18 (1.15-1.21; p<0.0001) |
| Antihypertensive drugs | 3.11 (2.77-3.50; p<0.0001) | 2.61 (2.31-2.95; p<0.0001) |
| Lipid-lowering drugs | 2.19 (1.99-2.41; p<0.0001) | 1.95 (1.76-2.16; p<0.0001) |
| Anticoagulants | 4.01 (3.14-5.12; p<0.0001) | 3.22 (2.52-4.12; p<0.0001) |
| Antiplatelets | 3.01 (2.74-3.31; p<0.0001) | 2.54 (2.29-2.81; p<0.0001) |
| ***Laboratory tests*** |  |  |
| Lymphocyte, x10^9/L | 0.69 (0.67-0.72; p<0.0001) | 0.74 (0.71-0.77; p<0.0001) |
| Neutrophil, x10^9/L | 1.065 (1.056-1.074; p<0.0001) | 1.05 (1.04-1.06; p<0.0001) |
| Platelet, x10^9/L | 0.999 (0.999-1.000; p=0.0052) | 0.999 (0.998-1.000; p=0.0045) |
| Potassium, mmol/L | 0.98 (0.95-1.01; p=0.1232) | 1.05 (1.01-1.09; p=0.0177) |
| Albumin, g/L | 0.901 (0.897-0.904; p<0.0001) | 0.91 (0.90-0.91; p<0.0001) |
| Sodium, mmol/L | 0.96 (0.96-0.97; p<0.0001) | 0.95 (0.95-0.96; p<0.0001) |

| Urea, mmol/L | 1.087 (1.085-1.090; p<0.0001) | 1.07 (1.07-1.08; p<0.0001) |
| --- | --- | --- |
| Creatinine, umol/L | 1.002 (1.002-1.002; p<0.0001) | 1.002 (1.002-1.002; p<0.0001) |
| Total protein, g/L | 0.97 (0.97-0.97; p<0.0001) | 0.97 (0.97-0.97; p<0.0001) |
| Alkaline phosphatase, U/L | 1.003 (1.003-1.003; p<0.0001) | 1.003 (1.003-1.003; p<0.0001) |
| Aspartate transaminase, U/L | 1.001 (1.000-1.001; p=0.0307) | 1.000 (0.999-1.001; p=0.4263) |
| Alanine transaminase, U/L | 0.988 (0.987-0.990; p<0.0001) | 0.988 (0.986-0.989; p<0.0001) |
| Bilirubin, umol/L | 1.002 (0.996-1.008; p=0.5319) | 1.002 (0.996-1.008; p=0.5399) |
| Triglyceride, mmol/L | 0.99 (0.98-1.00; p=0.0470) | 0.97 (0.96-0.99; p=0.0001) |
| Low-density lipoprotein, mmol/L | 0.90 (0.88-0.91; p<0.0001) | 0.86 (0.84-0.88; p<0.0001) |
| Total cholesterol, mmol/L | 0.89 (0.88-0.90; p<0.0001) | 0.86 (0.84-0.87; p<0.0001) |
| High-density lipoprotein, mmol/L | 0.83 (0.80-0.86; p<0.0001) | 0.79 (0.75-0.83; p<0.0001) |
| HbA1c, g/dL | 0.77 (0.76-0.78; p<0.0001) | 0.80 (0.79-0.81; p<0.0001) |
| Glucose, mmol/L | 1.02 (1.02-1.03; p<0.0001) | 1.03 (1.03-1.04; p<0.0001) |

CAIDE: cardiovascular risk factors, aging, and incidence of dementia, AMI: acute myocardial infarction, COPD: chronic obstructive pulmonary disease, IHD: ischemic heart disease, PVD: peripheral vascular disease, TIA: transient ischemic attack, ACEI: angiotensin-converting-enzyme inhibitors, ARB: angiotensin II receptor blockers, LDL: low density lipoprotein cholesterol, HDL: high density lipoprotein cholesterol, HR: hazard ratio, CI: confidence interval.
