## Supplementary Table 1 for "Metformin use is associated with lower risks of dementia, anxiety and depression: The Hong Kong Diabetes Study"

Supplementary Table 1. ICD-9 codes for disease diagnoses.

Diabetes mellitus 250	250.01 250.02 250.03 250.1	250.11 250.12 250.13 250.2	250.21

250.22 250.23 250.3	250.31 250.32 250.33 250.4	250.41 250.42 250.43 250.5

250.51 250.52 250.53 250.6	250.61 250.62 250.63 250.7	250.71 250.72 250.73

250.8	250.81 250.82 250.83 250.9	250.91 250.92 250.93

Renal diseases 582	582	582.1	582.2	582.4	582.8	582.81 582.89 582.9	583

583	583.1	583.2	583.4	583.6	583.7	585	585.1	585.2	585.3	585.4

585.5	585.6	585.9	586	588	588	588.1	588.8	588.81 588.89 588.9

Coronary heart disease 414

Hypertension	401	401.1	401.9	402	402.01 402.1	402.11 402.9	402.91 403

403.01 403.1	403.11 403.9	403.91 404	404.01 404.02 404.03 404.1	404.11

404.12 404.13 404.9	404.91 404.92 404.93 405	405.01 405.09 405.1	405.11

405.19 405.9	405.91 405.99 437.2

Heart failure	428	428	428.1	428.2	428.2	428.21 428.22 428.23 428.3	428.3

428.31 428.32 428.33 428.4	428.4	428.41 428.42 428.43 428.9	398.91 402.01

402.11 402.91 404.01 404.03 404.11 404.13 404.91 404.93

Atrial fibrillation	427.31 429.4

Hemiplegia or paraplegia 344.1 342	342	342	342.01 342.02 342.1	342.1	342.11

342.12 342.8	342.8	342.81 342.82 342.9	342.9	342.91 342.92

Osteoporosis 733.00 733.01 733.02 733.03 733.09 733.41 733.42 733.43 733.40 731.0 733.7

733.00

Neurologic 250.6	250.61 251.61 252.61

Anemia 281 282 283 285

Ventricular tachycardia/fibrillation 427.41 427.1

Ophthalmic 250.5	250.51 250.52 250.53

Liver diseases	456	456.1	456.2	572.2	572.3	572.4	572.8	571.4	571.5	571.6

Dementia and Alzheimer	331.82 290	290.1	290.11 290.12 290.13 290.2	290.21

290.3	290.4	290.41 290.42 290.43 290.8	290.9	294.2	294.1	294.11 294.21

332	46.1	333.4	340	42331 331.19 294.29

Anxiety disorder 300.0 300.01 300.02 300.2 300.21 300.23 300.3

COPD 490	491	492	493	494	495	496	491.1	491.2	491.21 491.22

491.8	491.9	492.8	493.01 493.02 493.1	493.11 493.12 493.2	493.21 493.22

493.8	493.81 493.82 493.9	493.91 493.92 494.1	495.1	495.2	495.3	495.4

495.5	495.6	495.7	495.8	495.9

PVD	250.7	443.9	443	443.1	443.2	443.21 443.22 443.23 443.24 443.29 443.8

443.81 443.82 443.89 441	443.9	785.4	V43.4

Stroke/TIA	Stroke/TIA	435	435.1	435.2	435.3	435.8	435.9	433.81 433.91

434	436	437	437.1	433.31 433.01 434.01 434.1	434.11 434.9	434.91

437.2	437.3	437.4	437.5	437.6	437.7	437.8	437.9	430	431	432

432.1	432.9

Gastrointestinal bleeding	531	531.2	531.4	531.6	532	532.2	532.4	532.6

533	533.2	533.4	533.6	534	534.2	534.4	534.6	535.01 535.11 535.21

535.31 535.41 535.51 535.61 535.71 562.02 562.03 562.12 562.13 569.3	569.85

569.86 578	578.1	578.9

IHD	410.01 410.02 410.1	410.11 410.12 410.2	410.21 410.22 410.3	410.31 410.32

410.4	410.41 410.42 410.5	410.51 410.52 410.6	410.61 410.62 410.7	410.71

410.72 410.8	410.81 410.82 410.9	410.91 410.92 411	411.1	411.8	411.81

411.89 413	413.1	413.9	414	414.01 414.02 414.03 414.04 414.05 414.06

414.07 414.1	414.11 414.12 414.19 414.2	414.3	414.4	414.8	414.9	410

412

| Malignancy 140140.1 | | 140.3 | 140.4 | 140.5 | 140.6 | 140.8 | 140.9 | 141 | 141.1 | 141.2 |
| --- | --- | --- | --- | --- | --- | --- | --- | --- | --- | --- |
| 141.3 | 141.4 | 141.5 | 141.6 | 141.8 | 141.9 | 142 | 142.1 | 142.2 | 142.8 | 142.9 |
| 143 | 143.1 | 143.8 | 143.9 | 144 | 144.1 | 144.8 | 144.9 | 145 | 145.1 | 145.2 |
| 145.3 | 145.4 | 145.5 | 145.6 | 145.8 | 145.9 | 146 | 146.1 | 146.2 | 146.3 | 146.4 |
| 146.5 | 146.6 | 146.7 | 146.8 | 146.9 | 147 | 147.1 | 147.2 | 147.3 | 147.8 | 147.9 |
| 148 | 148.1 | 148.2 | 148.3 | 148.8 | 148.9 | 149 | 149.1 | 149.8 | 149.9 | 150 |
| 150.1 | 150.2 | 150.3 | 150.4 | 150.5 | 150.8 | 150.9 | 151 | 151.1 | 151.2 | 151.3 |
| 151.4 | 151.5 | 151.6 | 151.8 | 151.9 | 152 | 152.1 | 152.2 | 152.3 | 152.8 | 152.9 |
| 153 | 153.1 | 153.2 | 153.3 | 153.4 | 153.5 | 153.6 | 153.7 | 153.8 | 153.9 | 154 |
| 154.1 | 154.2 | 154.3 | 154.8 | 155 | 155.1 | 155.2 | 156 | 156.1 | 156.2 | 156.8 |
| 156.9 | 157 | 157.1 | 157.2 | 157.3 | 157.4 | 157.8 | 157.9 | 158 | 158.8 | 158.9 |
| 159 | 159.1 | 159.8 | 159.9 | 160 | 160.1 | 160.2 | 160.3 | 160.4 | 160.5 | 160.8 |
| 160.9 | 161 | 161.1 | 161.2 | 161.3 | 161.8 | 161.9 | 162 | 162.2 | 162.3 | 162.4 |
| 162.5 | 162.8 | 162.9 | 163 | 163.1 | 163.8 | 163.9 | 164 | 164.1 | 164.2 | 164.3 |
| 164.8 | 164.9 | 165 | 165.8 | 165.9 | 170 | 170.1 | 170.2 | 170.3 | 170.4 | 170.5 |
| 170.6 | 170.7 | 170.8 | 170.9 | 171 | 171.2 | 171.3 | 171.4 | 171.5 | 171.6 | 171.7 |
| 171.8 | 171.9 | 172 | 172.1 | 172.2 | 172.3 | 172.4 | 172.5 | 172.6 | 172.7 | 172.8 |
| 172.9 | and | 173 | 173.01 | 173.02 | 173.09 | 173.1 | 173.11 | 173.12 | 173.19 | 173.2 |
| 173.21 | 173.22 | 173.29 | 173.3 | 173.31 | 173.32 | 173.39 | 173.4 | 173.41 | 173.42 | 173.49 |
| 173.5 | 173.51 | 173.52 | 173.59 | 173.6 | 173.61 | 173.62 | 173.69 | 173.7 | 173.71 | 173.72 |
| 173.79 | 173.8 | 173.81 | 173.82 | 173.89 | 173.9 | 173.91 | 173.92 | 173.99 | 174 | 174.1 |
| 174.2 | 174.3 | 174.4 | 174.5 | 174.6 | 174.8 | 174.9 | 175 | 175.9 | 176 | 176.1 |
| 176.2 | 176.3 | 176.4 | 176.5 | 176.8 | 176.9 | 179 | 180 | 180.1 | 180.8 | 180.9 |
| 181 | 182 | 182.1 | 182.8 | 183 | 183.2 | 183.3 | 183.4 | 183.5 | 183.8 | 183.9 |
| 184 | 184.1 | 184.2 | 184.3 | 184.4 | 184.8 | 184.9 | 185 | 186 | 186.9 | 187 |
| 187.1 | 187.2 | 187.3 | 187.4 | 187.5 | 187.6 | 187.7 | 187.8 | 187.9 | 188 | 188.1 |
| 188.2 | 188.3 | 188.4 | 188.5 | 188.6 | 188.7 | 188.8 | 188.9 | 189 | 189.1 | 189.2 |
| 189.3 | 189.4 | 189.8 | 189.9 | 190 | 190.1 | 190.2 | 190.3 | 190.4 | 190.5 | 190.6 |
| 190.7 | 190.8 | 190.9 | 191 | 191.1 | 191.2 | 191.3 | 191.4 | 191.5 | 191.6 | 191.7 |
| 191.8 | 191.9 | 192 | 192.1 | 192.2 | 192.3 | 192.8 | 192.9 | 193 | 194 | 194.1 |
| 194.3 | 194.4 | 194.5 | 194.6 | 194.8 | 194.9 | 195 | 195.1 | 195.2 | 195.3 | 195.4 |
| 195.5 | 195.8 | 200 | 200.01 | 200.02 | 200.03 | 200.04 | 200.05 | 200.06 | 200.07 | 200.08 |
| 200.1 | 200.11 | 200.12 | 200.13 | 200.14 | 200.15 | 200.16 | 200.17 | 200.18 | 200.2 | 200.21 |
| 200.22 | 200.23 | 200.24 | 200.25 | 200.26 | 200.27 | 200.28 | 200.3 | 200.31 | 200.32 | 200.33 |
| 200.34 | 200.35 | 200.36 | 200.37 | 200.38 | 200.4 | 200.41 | 200.42 | 200.43 | 200.44 | 200.45 |
| 200.46 | 200.47 | 200.48 | 200.5 | 200.51 | 200.52 | 200.53 | 200.54 | 200.55 | 200.56 | 200.57 |
| 200.58 | 200.6 | 200.61 | 200.62 | 200.63 | 200.64 | 200.65 | 200.66 | 200.67 | 200.68 | 200.7 |
| 200.71 | 200.72 | 200.73 | 200.74 | 200.75 | 200.76 | 200.77 | 200.78 | 200.8 | 200.81 | 200.82 |
| 200.83 | 200.84 | 200.85 | 200.86 | 200.87 | 200.88 | 201 | 201.01 | 201.02 | 201.03 | 201.04 |
| 201.05 | 201.06 | 201.07 | 201.08 | 201.1 | 201.11 | 201.12 | 201.13 | 201.14 | 201.15 | 201.16 |
| 201.17 | 201.18 | 201.2 | 201.21 | 201.22 | 201.23 | 201.24 | 201.25 | 201.26 | 201.27 | 201.28 |
| 201.4 | 201.41 | 201.42 | 201.43 | 201.44 | 201.45 | 201.46 | 201.47 | 201.48 | 201.5 | 201.51 |
| 201.52 | 201.53 | 201.54 | 201.55 | 201.56 | 201.57 | 201.58 | 201.6 | 201.61 | 201.62 | 201.63 |
| 201.64 | 201.65 | 201.66 | 201.67 | 201.68 | 201.7 | 201.71 | 201.72 | 201.73 | 201.74 | 201.75 |
| 201.76 | 201.77 | 201.78 | 201.9 | 201.91 | 201.92 | 201.93 | 201.94 | 201.95 | 201.96 | 201.97 |
| 201.98 | 202 | 202.01 | 202.02 | 202.03 | 202.04 | 202.05 | 202.06 | 202.07 | 202.08 | 202.1 |
| 202.11 | 202.12 | 202.13 | 202.14 | 202.15 | 202.16 | 202.17 | 202.18 | 202.2 | 202.21 | 202.22 |
| 202.23 | 202.24 | 202.25 | 202.26 | 202.27 | 202.28 | 202.3 | 202.31 | 202.32 | 202.33 | 202.34 |
| 202.35 | 202.36 | 202.37 | 202.38 | 202.4 | 202.41 | 202.42 | 202.43 | 202.44 | 202.45 | 202.46 |
| 202.47 | 202.48 | 202.5 | 202.51 | 202.52 | 202.53 | 202.54 | 202.55 | 202.56 | 202.57 | 202.58 |
| 202.6 | 202.61 | 202.62 | 202.63 | 202.64 | 202.65 | 202.66 | 202.67 | 202.68 | 202.7 | 202.71 |

202.72 202.73 202.74 202.75 202.76 202.77 202.78 202.8	202.81 202.82 202.83

202.84 202.85 202.86 202.87 202.88 202.9	202.91 202.92 202.93 202.94 202.95

202.96 202.97 202.98 203	203.01 203.02 203.1	203.11 203.12 203.8	203.81

203.82 204	204.01 204.02 204.1	204.11 204.12 204.2	204.21 204.22 204.8

204.81 204.82 204.9	204.91 204.92 205	205.01 205.02 205.1	205.11 205.12

205.2	205.21 205.22 205.3	205.31 205.32 205.8	205.81 205.82 205.9	205.91

205.92 206	206.01 206.02 206.1	206.11 206.12 206.2	206.21 206.22 206.8

206.81 206.82 206.9	206.91 206.92 207	207.01 207.02 207.1	207.11 207.12

207.2	207.21 207.22 207.8	207.81 207.82 208	208.01 208.02 208.1	208.11

208.12 208.2	208.21 208.22 208.8	208.81 208.82 208.9	208.91 208.92

Metastatic solid tumor 196	196.1	196.2	196.3	196.5	196.6	196.8	196.9	197

197.1	197.2	197.3	197.4	197.5	197.6	197.7	197.8	198	198.1	198.2

198.3	198.4	198.5	198.6	198.7	198.8	198.81 198.82 198.89 199	199.1

Obesity 278.01 278	278
