## Supplementary Table 2 for "Metformin use is associated with lower risks of dementia, anxiety and depression: The Hong Kong Diabetes Study"

Supplementary Table 2. Probability regression analysis of confounding characteristics to predict all-cause mortality before conducting propensity score matching.

| **Confounding characteristics** | **Statistics** |  |  |  | **[95% Conf. Interval]** | |
| --- | --- | --- | --- | --- | --- | --- |
|  | **Coef.** | **Std. Err.** | **t** | **P>t** | **Lower** | **Upper** |
| Metformin v.s. sulphonylurea | -0.108 | 0.007 | -14.89 | <0.001 | -0.122 | -0.093 |
| Male gender | 0.156 | 0.011 | 14.2 | <0.001 | 0.134 | 0.177 |
| Baseline age, years | 0.009 | 0.001 | 10.56 | <0.001 | 0.008 | 0.011 |
| CAIDE score | -0.016 | 0.004 | -3.93 | <0.001 | -0.024 | -0.008 |
| CHA-DS-VASc score | 0.077 | 0.009 | 9.06 | <0.001 | 0.061 | 0.094 |
| Charlson comorbidity index | 0.025 | 0.005 | 4.55 | <0.001 | 0.014 | 0.035 |
| Number of comorbidities | -0.027 | 0.007 | -3.86 | <0.001 | -0.040 | -0.013 |
| Hypertension | 0.039 | 0.015 | 2.65 | 0.008 | 0.010 | 0.067 |
| Heart failure | 0.012 | 0.016 | 0.71 | 0.48 | -0.021 | 0.044 |
| Coronary heart disease | 0.047 | 0.013 | 3.68 | <0.001 | 0.022 | 0.073 |
| Atrial fibrillation | 0.090 | 0.020 | 4.52 | <0.001 | 0.051 | 0.129 |
| Hemiplegia or paraplegia | -0.028 | 0.029 | -0.96 | 0.335 | -0.084 | 0.029 |
| Renal diseases | 0.107 | 0.017 | 6.45 | <0.001 | 0.074 | 0.140 |
| Neurologic | 0.085 | 0.019 | 4.46 | <0.001 | 0.048 | 0.123 |
| Ophthalmic | 0.119 | 0.013 | 8.87 | <0.001 | 0.093 | 0.145 |
| Liver diseases | 0.170 | 0.038 | 4.42 | <0.001 | 0.095 | 0.246 |
| Ventricular tachycardia/fibrillation | 0.065 | 0.038 | 1.73 | 0.084 | -0.009 | 0.138 |
| Anemia | 0.196 | 0.011 | 18.33 | <0.001 | 0.175 | 0.217 |
| AMI | -0.041 | 0.028 | -1.47 | 0.142 | -0.097 | 0.014 |
| COPD | 0.051 | 0.022 | 2.34 | 0.019 | 0.008 | 0.094 |
| IHD | -0.040 | 0.016 | -2.47 | 0.013 | -0.072 | -0.008 |
| PVD | 0.041 | 0.026 | 1.59 | 0.113 | -0.010 | 0.092 |
| Stroke/TIA | -0.129 | 0.021 | -6.28 | <0.001 | -0.170 | -0.089 |
| Gastrointestinal bleeding | 0.033 | 0.020 | 1.67 | 0.094 | -0.006 | 0.072 |

| Obesity | -0.007 | 0.022 | -0.32 | 0.747 | -0.051 | 0.037 |
| --- | --- | --- | --- | --- | --- | --- |
| Other anti-diabetic drugs | 0.012 | 0.008 | 1.61 | 0.108 | -0.003 | 0.028 |
| Total cholesterol, mmol/L | -0.003 | 0.000 | -6.88 | <0.001 | -0.004 | -0.002 |

CAIDE: cardiovascular risk factors, aging, and incidence of dementia, AMI: acute myocardial infarction, COPD: chronic obstructive pulmonary disease, IHD: ischemic heart disease, PVD: peripheral vascular disease, TIA: transient ischemic attack.
