## Supplementary Table 3 for "Metformin use is associated with lower risks of dementia, anxiety and depression: The Hong Kong Diabetes Study"

Supplementary Table 3. Confounding balancing comparisons of metformin and sulphonylurea after propensity score matching with 1:1 nearest neighbor search using Stata.

| **Confounding characteristics** | **Mean** |  |  | **t-test** |  | **V(T)/V(C)** |
| --- | --- | --- | --- | --- | --- | --- |
|  | **Treated** | **Control** | **%bias** | **t** | **p>t** |  |
| Male gender | 0.449 | 0.464 | -2.9 | -2.07 | 0.038 | . |
| Baseline age, years | 65.200 | 65.186 | 0.1 | 0.08 | 0.939 | 0.82* |
| CAIDE score | 4.534 | 4.509 | 1.5 | 1.04 | 0.299 | 0.82* |
| CHA-DS-VASc score | 2.406 | 2.441 | -2.1 | -1.49 | 0.136 | 0.91* |
| Charlson comorbidity index | 3.324 | 3.370 | -2 | -1.45 | 0.148 | 0.90* |
| Number of comorbidities | 2.378 | 2.434 | -2.3 | -1.68 | 0.094 | 0.92* |
| Hypertension | 0.375 | 0.376 | -0.2 | -0.13 | 0.897 | . |
| Heart failure | 0.061 | 0.071 | -4 | -2.91 | 0.004 | . |
| Coronary heart disease | 0.123 | 0.118 | 1.2 | 0.98 | 0.325 | . |
| Atrial fibrillation | 0.036 | 0.037 | -0.8 | -0.6 | 0.551 | . |
| Hemiplegia or paraplegia | 0.019 | 0.020 | -1.4 | -0.9 | 0.366 | . |
| Renal diseases | 0.047 | 0.050 | -1.1 | -0.94 | 0.347 | . |
| Neurologic | 0.025 | 0.022 | 1.5 | 1.47 | 0.142 | . |
| Ophthalmic | 0.088 | 0.092 | -1.3 | -1.05 | 0.294 | . |
| Liver diseases | 0.006 | 0.011 | -6.1 | -4.3 | 0 | . |
| Ventricular tachycardia/fibrillation | 0.008 | 0.005 | 2.4 | 1.91 | 0.057 | . |
| Anemia | 0.175 | 0.180 | -1 | -0.82 | 0.412 | . |
| AMI | 0.020 | 0.023 | -2.5 | -1.6 | 0.111 | . |
| COPD | 0.027 | 0.032 | -3.7 | -2.43 | 0.015 | . |
| IHD | 0.111 | 0.112 | -0.4 | -0.27 | 0.791 | . |
| PVD | 0.016 | 0.021 | -3.8 | -2.67 | 0.008 | . |
| Stroke/TIA | 0.076 | 0.078 | -0.8 | -0.57 | 0.566 | . |
| Gastrointestinal bleeding | 0.032 | 0.036 | -2.7 | -1.88 | 0.06 | . |
| Obesity | 0.039 | 0.033 | 3.3 | 1.98 | 0.048 | . |

| Other anti-diabetic drugs | 0.300 | 0.324 | -5.7 | -3.82 | 0 | . |
| --- | --- | --- | --- | --- | --- | --- |
| Total cholesterol, mmol/L | 74.703 | 74.911 | -3 | -2.25 | 0.025 | 0.81* |

CAIDE: cardiovascular risk factors, aging, and incidence of dementia, AMI: acute myocardial infarction, COPD: chronic obstructive pulmonary disease, IHD: ischemic heart disease, PVD: peripheral vascular disease, TIA: transient ischemic attack
