## Supplementary Table 4 for "Metformin use is associated with lower risks of dementia, anxiety and depression: The Hong Kong Diabetes Study"

Supplementary Table 4. Estimations of bootstrapped standard error (replications=50) for the propensity matching with 1:1 nearest neighbor search using Stata.

| **Observed Coef.** | **Bootstrap Std. Err.** | **z** | **P>z** | **Normal based [95% Conf. Interval]** |
| --- | --- | --- | --- | --- |
| -0.107 | 0.012 | -8.7 | <0.001 | [-0.131, -0.083] |
