## Supplementary Table 5 for "Metformin use is associated with lower risks of dementia, anxiety and depression: The Hong Kong Diabetes Study"

Supplementary Table 5. Baseline and clinical characteristics of patients with type 2 diabetes mellitus by new diagnosis of dementia before and after 1:1 propensity score matching.

* for SMD≥0.2

| **Before matching Characteristics Dementia (N=3401)** | **No dementia (N=86370)** | **SMD** | **After 1:1 matching Dementia (N=1805)** | **No dementia (N=40683)** | **SMD** |
| --- | --- | --- | --- | --- | --- |
| **Mean(SD);N or Count(%)** | **Mean(SD);N or Count(%)** |  | **Mean(SD);N or Count(%)** | **Mean(SD);N or Count(%)** |  |
| ***Demographics***  Male gender 1132(33.28%) | 39826(46.11%) | 0.26* | 639(35.40%) | 19515(47.96%) | 0.26* |
| Female gender 2269(66.71%) | 46544(53.88%) | 0.26* | 1166(64.59%) | 21168(52.03%) | 0.26* |
| Baseline age, 76.4(7.8);n=3401 | 66.1(12.5);n=86370 | 0.99* | 77.0(7.9);n=1805 | 66.9(12.9);n=40683 | 0.94* |
| <50 10(0.29%) | 9356(10.83%) | 0.47* | 7(0.38%) | 4336(10.65%) | 0.46* |
| [50, 60) 96(2.82%) | 18178(21.04%) | 0.59* | 56(3.10%) | 7813(19.20%) | 0.53* |
| [60, 70) 490(14.40%) | 21627(25.03%) | 0.27* | 225(12.46%) | 9466(23.26%) | 0.28* |
| [70, 80) 1605(47.19%) | 25319(29.31%) | 0.37* | 815(45.15%) | 12293(30.21%) | 0.31* |
| >=80 1200(35.28%) | 11890(13.76%) | 0.52* | 702(38.89%) | 6775(16.65%) | 0.51* |
| ***Past comorbidities*** |  |  |  |  |  |
| CAIDE score 4.8(1.1);n=3401 | 4.3(1.5);n=86370 | 0.4* | 4.9(1.1);n=1805 | 4.4(1.6);n=40683 | 0.37* |
| CHA-DS-VASc 3.2(1.5);n=3401 | 2.1(1.6);n=86370 | 0.69* | 3.4(1.5);n=1805 | 2.3(1.7);n=40683 | 0.67* |
| Charlson  comorbidity 4.1(1.7);n=3401 index | 3.0(2.0);n=86370 | 0.62* | 4.3(1.8);n=1805 | 3.1(2.1);n=40683 | 0.59* |
| Number of 1.9(2.1);n=3401 | 1.5(2.0);n=86370 | 0.2* | 2.3(2.3);n=1805 | 1.8(2.2);n=40683 | 0.2* |
| Hypertension 1241(36.48%) | 23393(27.08%) | 0.20* | 777(43.04%) | 13723(33.73%) | 0.19 |
| Heart failure 198(5.82%) | 3909(4.52%) | 0.06 | 140(7.75%) | 2399(5.89%) | 0.07 |
| Coronary heart 341(10.02%) | 7738(8.95%) | 0.04 | 236(13.07%) | 5117(12.57%) | 0.01 |
| Atrial 131(3.85%) | 2594(3.00%) | 0.05 | 86(4.76%) | 1549(3.80%) | 0.05 |

years

score

comorbidities

disease fibrillation

Hemiplegia or paraplegia Renal diseases Neurologic Ophthalmic Liver diseases Ventricular tachycardia/fibri llation

| 60(1.76%) | 1282(1.48%) | 0.02 | 41(2.27%) | 751(1.84%) | 0.03 |
| --- | --- | --- | --- | --- | --- |
| 110(3.23%) | 2945(3.40%) | 0.01 | 74(4.09%) | 1948(4.78%) | 0.03 |
| 22(0.64%) | 704(0.81%) | 0.02 | 21(1.16%) | 628(1.54%) | 0.03 |
| 57(1.67%) | 2130(2.46%) | 0.06 | 53(2.93%) | 1785(4.38%) | 0.08 |
| 11(0.32%) | 329(0.38%) | 0.01 | 9(0.49%) | 236(0.58%) | 0.01 |
| 26(0.76%) | 531(0.61%) | 0.02 | 18(0.99%) | 317(0.77%) | 0.02 |
| 135(3.96%) | 4919(5.69%) | 0.08 | 122(6.75%) | 4313(10.60%) | 0.14 |
| 66(1.94%) | 1263(1.46%) | 0.04 | 42(2.32%) | 625(1.53%) | 0.06 |
| 100(2.94%) | 1940(2.24%) | 0.04 | 69(3.82%) | 1117(2.74%) | 0.06 |
| 294(8.64%) | 6733(7.79%) | 0.03 | 192(10.63%) | 3807(9.35%) | 0.04 |
| 35(1.02%) | 879(1.01%) | 0 | 20(1.10%) | 579(1.42%) | 0.03 |
| 258(7.58%) | 4978(5.76%) | 0.07 | 161(8.91%) | 2799(6.88%) | 0.08 |
| 108(3.17%) | 2367(2.74%) | 0.03 | 81(4.48%) | 1475(3.62%) | 0.04 |
| 17(0.49%) | 533(0.61%) | 0.02 | 12(0.66%) | 304(0.74%) | 0.01 |
| 2(0.05%) | 119(0.13%) | 0.03 | 2(0.11%) | 59(0.14%) | 0.01 |
| 20(0.58%) | 1063(1.23%) | 0.07 | 13(0.72%) | 417(1.02%) | 0.03 |
| 2523(74.18%) | 66004(76.42%) | 0.05 | 927(51.35%) | 20317(49.93%) | 0.03 |
| 729(21.43%) | 24391(28.24%) | 0.16 | 352(19.50%) | 11909(29.27%) | 0.23* |
| 1719(50.54%) | 42548(49.26%) | 0.03 | 913(50.58%) | 18544(45.58%) | 0.1 |
| 1219(35.84%) | 29823(34.52%) | 0.03 | 677(37.50%) | 15000(36.87%) | 0.01 |
| 1666(48.98%) | 36848(42.66%) | 0.13 | 862(47.75%) | 17556(43.15%) | 0.09 |
| 781(22.96%) | 16618(19.24%) | 0.09 | 444(24.59%) | 9542(23.45%) | 0.03 |
| 742(21.81%) | 21618(25.02%) | 0.08 | 446(24.70%) | 10844(26.65%) | 0.04 |
| 18(0.52%) | 265(0.30%) | 0.03 | 16(0.88%) | 240(0.58%) | 0.03 |

Anemia AMI COPD IHD PVD

Stroke/TIA Gastrointestinal bleeding Malignancy Metastatic solid tumor

Obesity ***Medications*** Metformin v.s. sulphonylurea Other anti- diabetic drugs ACEI/ARB

Beta blockers Calcium channel blockers Diuretics

Stains Nitrates

Sulfadiazine Antihypertensiv e drugs

| 2454(72.15%) | 58185(67.36%) | 0.1 | 1114(61.71%) | 20398(50.13%) | 0.23* |
| --- | --- | --- | --- | --- | --- |
| 22(0.64%) | 346(0.40%) | 0.03 | 22(1.21%) | 318(0.78%) | 0.04 |
| 28(0.82%) | 657(0.76%) | 0.01 | 26(1.44%) | 562(1.38%) | 0.01 |
| 4(0.11%) | 75(0.08%) | 0.01 | 4(0.22%) | 75(0.18%) | 0.01 |
| 41(1.20%) | 543(0.62%) | 0.06 | 37(2.04%) | 475(1.16%) | 0.07 |
| 1.7(0.7);n=226 | 1.9(1.4);n=9719 | 0.12 | 1.7(0.7);n=180 | 1.8(1.5);n=7131 | 0.09 |
| 5.6(2.3);n=226 | 5.5(2.7);n=9719 | 0.04 | 5.5(2.3);n=180 | 5.6(2.8);n=7131 | 0.05 |
| 251.2(77.6);n=341 | 260.1(85.3);n=5381 | 0.11 | 255.5(74.7);n=188 | 256.4(83.5);n=2304 | 0.01 |
| 4.2(0.5);n=2013 | 4.3(0.5);n=48388 | 0.13 | 4.2(0.5);n=1113 | 4.2(0.5);n=22146 | 0.09 |
| 36.9(5.2);n=466 | 38.7(5.2);n=16138 | 0.35* | 36.4(5.5);n=306 | 38.1(5.5);n=9905 | 0.3* |
| 139.3(3.5);n=2013 | 139.3(3.1);n=48415 | 0 | 139.1(3.7);n=1113 | 139.3(3.3);n=22164 | 0.08 |
| 6.9(3.2);n=1755 | 6.8(4.0);n=43275 | 0.02 | 7.2(3.6);n=977 | 7.7(5.1);n=20256 | 0.11 |
| 97.3(63.5);n=1766 | 103.4(98.4);n=43511 | 0.07 | 105.2(81.2);n=985 | 124.2(138.0);n=20371 | 0.17 |
| 73.7(6.9);n=464 | 74.4(6.8);n=16088 | 0.11 | 73.4(7.3);n=305 | 74.1(7.2);n=9868 | 0.1 |
| 78.8(30.5);n=1773 | 78.0(35.3);n=40177 | 0.03 | 81.6(34.4);n=996 | 81.7(38.5);n=19066 | <0.01 |
| 34.5(122.9);n=195 | 31.7(64.7);n=3112 | 0.03 | 25.3(15.0);n=99 | 33.8(65.4);n=1612 | 0.18 |
| 20.1(15.8);n=1509 | 25.4(23.4);n=31986 | 0.27* | 20.5(17.0);n=867 | 25.8(23.4);n=15674 | 0.26* |

Lipid-lowering drugs Anticoagulants Antiplatelets ***Laboratory tests*** Lymphocyte, x10^9/L Neutrophil, x10^9/L Platelet, x10^9/L Potassium, mmol/L Albumin, g/L Sodium, mmol/L

Urea, mmol/L Creatinine, umol/L

Total protein,

g/L Alkaline

phosphatase, U/L Aspartate transaminase, U/L

Alanine transaminase, U/L

umol/L mmol/L

| Bilirubin, - | 13.4(11.5);n=1990 | - | - | 13.4(11.5);n=1990 | - |
| --- | --- | --- | --- | --- | --- |
| Triglyceride, 1.6(1.0);n=2301 | 1.7(1.4);n=62755 | 0.11 | 1.6(1.0);n=1270 | 1.8(1.4);n=29560 | 0.11 |
| Low-density  lipoprotein, 2.9(0.8);n=1327 | 2.9(0.9);n=39765 | 0.05 | 2.9(0.9);n=712 | 2.9(0.9);n=18488 | 0.08 |
| mmol/L  Total  cholesterol, 4.8(1.0);n=2314 | 4.8(1.0);n=62892 | 0.06 | 4.8(1.0);n=1277 | 4.9(1.1);n=29640 | 0.1 |
| mmol/L  High-density  lipoprotein, 1.2(0.4);n=2006 | 1.2(0.3);n=55643 | 0.1 | 1.2(0.4);n=1103 | 1.2(0.4);n=25474 | 0.11 |
| mmol/L |  |  |  |  |  |
| HbA1c, g/dL 11.9(1.6);n=234 | 12.5(2.0);n=11003 | 0.33* | 11.6(1.6);n=185 | 12.2(2.1);n=7963 | 0.3* |
| Glucose, 9.0(4.2);n=728 | 8.9(4.2);n=28611 | 0.04 | 8.5(3.6);n=508 | 8.5(4.1);n=20343 | 0.02 |

mmol/L

CAIDE: cardiovascular risk factors, aging, and incidence of dementia, AMI: acute myocardial infarction, COPD: chronic obstructive pulmonary disease, IHD: ischemic heart disease, PVD: peripheral vascular disease, TIA: transient ischemic attack, ACEI: angiotensin-converting-enzyme inhibitors, ARB: angiotensin II receptor blockers, LDL: low density lipoprotein cholesterol, HDL: high density lipoprotein cholesterol, SD: standard deviation, SMD: standard mean difference.
