## Supplementary Table 6 for "Metformin use is associated with lower risks of dementia, anxiety and depression: The Hong Kong Diabetes Study"

Supplementary Table 6. Baseline and clinical characteristics of patients with type 2 diabetes by new diagnosis of anxiety/ depression before and after 1:1 propensity score matching.

* for SMD≥0.2

|  | **Before matching** |  |  | **After 1:1 matching** |  |  |
| --- | --- | --- | --- | --- | --- | --- |
| **Characteristics** | **Anxiety disorder and depression (N=1493) Mean(SD);N or Count(%)** | **No anxiety disorder and depression (N=88278) Mean(SD);N or Count(%)** | **SMD** | **Anxiety disorder and depression (N=752) Mean(SD);N or Count(%)** | **No anxiety disorder and depression (N=41736) Mean(SD);N or Count(%)** | **SMD** |
| ***Demographics*** |  |  |  |  |  |  |
| Male gender | 438(29.33%) | 40520(45.90%) | 0.35* | 235(31.25%) | 19919(47.72%) | 0.34* |
| Female gender | 1055(70.66%) | 47758(54.09%) | 0.35* | 517(68.75%) | 21817(52.27%) | 0.34* |
| Baseline age, years | 63.9(12.9);n=1493 | 66.5(12.5);n=88278 | 0.2* | 64.7(13.2);n=752 | 67.4(12.9);n=41736 | 0.21* |
| <50 | 209(13.99%) | 9157(10.37%) | 0.11 | 109(14.49%) | 4234(10.14%) | 0.13 |
| [50, 60) | 375(25.11%) | 17899(20.27%) | 0.12 | 172(22.87%) | 7697(18.44%) | 0.11 |
| [60, 70) | 356(23.84%) | 21761(24.65%) | 0.02 | 164(21.80%) | 9527(22.82%) | 0.02 |
| [70, 80) | 392(26.25%) | 26532(30.05%) | 0.08 | 214(28.45%) | 12894(30.89%) | 0.05 |
| >=80 | 161(10.78%) | 12929(14.64%) | 0.12 | 93(12.36%) | 7384(17.69%) | 0.15 |
| ***Past comorbidities*** |  |  |  |  |  |  |
| CAIDE score | 4.3(1.7);n=1493 | 4.3(1.5);n=88278 | 0.02 | 4.4(1.7);n=752 | 4.5(1.5);n=41736 | 0.05 |
| CHA-DS-VASc  score | 2.2(1.5);n=1493 | 2.2(1.6);n=88278 | 0.01 | 2.3(1.6);n=752 | 2.3(1.7);n=41736 | 0.03 |
| Charlson comorbidity  index | 2.7(1.9);n=1493 | 3.0(2.0);n=88278 | 0.17 | 2.7(1.9);n=752 | 3.2(2.1);n=41736 | 0.25* |
| Number of comorbidities | 1.5(1.9);n=1493 | 1.6(2.0);n=88278 | 0.04 | 1.5(2.0);n=752 | 1.8(2.2);n=41736 | 0.14 |
| Hypertension | 458(30.67%) | 24176(27.38%) | 0.07 | 279(37.10%) | 14221(34.07%) | 0.06 |
| Heart failure | 57(3.81%) | 4050(4.58%) | 0.04 | 33(4.38%) | 2506(6.00%) | 0.07 |
| Coronary heart disease | 170(11.38%) | 7909(8.95%) | 0.08 | 116(15.42%) | 5237(12.54%) | 0.08 |
| Atrial fibrillation | 43(2.88%) | 2682(3.03%) | 0.01 | 21(2.79%) | 1614(3.86%) | 0.06 |

| Hemiplegia or  paraplegia | 22(1.47%) | 1320(1.49%) | 0 | 15(1.99%) | 777(1.86%) | 0.01 |
| --- | --- | --- | --- | --- | --- | --- |
| Renal diseases | 22(1.47%) | 3033(3.43%) | 0.13 | 14(1.86%) | 2008(4.81%) | 0.16 |
| Neurologic | 11(0.73%) | 715(0.80%) | 0.01 | 10(1.32%) | 639(1.53%) | 0.02 |
| Ophthalmic | 16(1.07%) | 2171(2.45%) | 0.11 | 15(1.99%) | 1823(4.36%) | 0.14 |
| Liver diseases | 2(0.13%) | 338(0.38%) | 0.05 | 2(0.26%) | 243(0.58%) | 0.05 |
| Ventricular tachycardia/fibri  llation | 6(0.40%) | 551(0.62%) | 0.03 | 2(0.26%) | 333(0.79%) | 0.07 |
| Anemia | 43(2.88%) | 5011(5.67%) | 0.14 | 33(4.38%) | 4402(10.54%) | 0.24* |
| AMI | 17(1.13%) | 1312(1.48%) | 0.03 | 10(1.32%) | 657(1.57%) | 0.02 |
| COPD | 39(2.61%) | 2001(2.26%) | 0.02 | 18(2.39%) | 1168(2.79%) | 0.03 |
| IHD | 113(7.56%) | 6914(7.83%) | 0.01 | 59(7.84%) | 3940(9.44%) | 0.06 |
| PVD | 9(0.60%) | 905(1.02%) | 0.05 | 4(0.53%) | 595(1.42%) | 0.09 |
| Stroke/TIA | 68(4.55%) | 5168(5.85%) | 0.06 | 34(4.52%) | 2926(7.01%) | 0.11 |
| Gastrointestinal bleeding | 33(2.21%) | 2442(2.76%) | 0.04 | 23(3.05%) | 1533(3.67%) | 0.03 |
| Malignancy | 1(0.06%) | 549(0.62%) | 0.09 | 1(0.13%) | 315(0.75%) | 0.09 |
| Metastatic solid tumor | 0(0.00%) | 121(0.13%) | 0.05 | 0(0.00%) | 61(0.14%) | 0.05 |
| Obesity | 14(0.93%) | 1069(1.21%) | 0.03 | 6(0.79%) | 424(1.01%) | 0.02 |
| ***Medications*** |  |  |  |  |  |  |
| Metformin v.s. sulphonylurea | 1041(69.72%) | 67486(76.44%) | 0.15 | 300(39.89%) | 20944(50.18%) | 0.21* |
| Other anti-  diabetic drugs | 407(27.26%) | 24713(27.99%) | 0.02 | 228(30.31%) | 12033(28.83%) | 0.03 |
| ACEI/ARB | 683(45.74%) | 43584(49.37%) | 0.07 | 317(42.15%) | 19140(45.85%) | 0.07 |
| Beta blockers | 565(37.84%) | 30477(34.52%) | 0.07 | 301(40.02%) | 15376(36.84%) | 0.07 |
| Calcium channel blockers | 565(37.84%) | 37949(42.98%) | 0.1 | 294(39.09%) | 18124(43.42%) | 0.09 |
| Diuretics | 278(18.62%) | 17121(19.39%) | 0.02 | 168(22.34%) | 9818(23.52%) | 0.03 |
| Stains | 421(28.19%) | 21939(24.85%) | 0.08 | 222(29.52%) | 11068(26.51%) | 0.07 |
| Nitrates | 5(0.33%) | 278(0.31%) | 0 | 4(0.53%) | 252(0.60%) | 0.01 |

| Sulfadiazine | 963(64.50%) | 59676(67.60%) | 0.07 | 363(48.27%) | 21149(50.67%) | 0.05 |
| --- | --- | --- | --- | --- | --- | --- |
| Antihypertensiv e drugs | 3(0.20%) | 365(0.41%) | 0.04 | 3(0.39%) | 337(0.80%) | 0.05 |
| Lipid-lowering drugs | 10(0.66%) | 675(0.76%) | 0.01 | 6(0.79%) | 582(1.39%) | 0.06 |
| Anticoagulants | 0(0.00%) | 79(0.08%) | 0.04 | 0(0.00%) | 79(0.18%) | 0.06 |
| Antiplatelets | 8(0.53%) | 576(0.65%) | 0.02 | 5(0.66%) | 507(1.21%) | 0.06 |
| ***Laboratory tests*** |  |  |  |  |  |  |
| Lymphocyte, x10^9/L | 2.1(0.8);n=105 | 1.9(1.4);n=9840 | 0.2* | 2.0(0.8);n=66 | 1.8(1.5);n=7245 | 0.15 |
| Neutrophil,  x10^9/L | 5.1(1.8);n=105 | 5.5(2.7);n=9840 | 0.19 | 5.2(1.8);n=66 | 5.6(2.8);n=7245 | 0.17 |
| Platelet,  x10^9/L | 271.5(82.6);n=157 | 259.2(84.9);n=5565 | 0.15 | 271.5(68.7);n=87 | 255.8(83.2);n=2405 | 0.21* |
| Potassium, mmol/L | 4.2(0.4);n=898 | 4.3(0.5);n=49503 | 0.16 | 4.2(0.4);n=454 | 4.2(0.5);n=22805 | 0.14 |
| Albumin, g/L | 38.5(4.8);n=229 | 38.6(5.2);n=16375 | 0.02 | 37.9(4.9);n=130 | 38.1(5.5);n=10081 | 0.03 |
| Sodium, mmol/L | 139.2(3.3);n=898 | 139.3(3.2);n=49530 | 0.04 | 139.3(3.3);n=454 | 139.3(3.3);n=22823 | 0.02 |
| Urea, mmol/L | 6.0(3.1);n=766 | 6.8(4.0);n=44264 | 0.24* | 6.3(3.8);n=386 | 7.7(5.1);n=20847 | 0.32* |
| Creatinine, umol/L | 88.7(70.6);n=770 | 103.4(97.7);n=44507 | 0.17 | 97.7(96.0);n=388 | 123.8(136.5);n=20968 | 0.22* |
| Total protein,  g/L | 74.0(6.5);n=229 | 74.4(6.8);n=16323 | 0.06 | 73.1(6.5);n=130 | 74.1(7.2);n=10043 | 0.15 |
| Alkaline  phosphatase, U/L | 77.7(26.1);n=850 | 78.0(35.3);n=41100 | 0.01 | 78.6(26.0);n=429 | 81.8(38.6);n=19633 | 0.1 |
| Aspartate  transaminase, U/L | 24.0(10.5);n=71 | 32.0(70.2);n=3236 | 0.16 | 25.5(10.8);n=35 | 33.5(64.2);n=1676 | 0.17 |
| Alanine transaminase,  U/L | 27.0(20.3);n=709 | 25.1(23.2);n=32786 | 0.09 | 27.2(18.7);n=367 | 25.5(23.2);n=16174 | 0.08 |

| Bilirubin,  umol/L | - | 13.4(11.5);n=1990 | - | - | 13.4(11.5);n=1990 | - |
| --- | --- | --- | --- | --- | --- | --- |
| Triglyceride, mmol/L | 1.9(2.5);n=1124 | 1.7(1.4);n=63932 | 0.1 | 1.8(1.2);n=563 | 1.7(1.4);n=30267 | 0.06 |
| Low-density  lipoprotein, mmol/L | 3.0(0.9);n=724 | 2.9(0.9);n=40368 | 0.11 | 3.0(0.9);n=357 | 2.9(0.9);n=18843 | 0.12 |
| Total cholesterol,  mmol/L | 5.0(1.1);n=1126 | 4.8(1.0);n=64080 | 0.12 | 5.0(1.0);n=564 | 4.9(1.1);n=30353 | 0.09 |
| High-density lipoprotein,  mmol/L | 1.2(0.4);n=997 | 1.2(0.3);n=56652 | 0.12 | 1.2(0.4);n=493 | 1.2(0.4);n=26084 | 0.11 |
| HbA1c, g/dL | 12.3(1.7);n=112 | 12.5(2.0);n=11125 | 0.08 | 11.9(1.8);n=71 | 12.2(2.1);n=8077 | 0.11 |
| Glucose, mmol/L | 8.4(4.5);n=555 | 8.9(4.2);n=28784 | 0.11 | 8.2(4.6);n=428 | 8.5(4.1);n=20423 | 0.07 |

CAIDE: cardiovascular risk factors, aging, and incidence of dementia, AMI: acute myocardial infarction, COPD: chronic obstructive pulmonary disease, IHD: ischemic heart disease, PVD: peripheral vascular disease, TIA: transient ischemic attack, ACEI: angiotensin-converting-enzyme inhibitors, ARB: angiotensin II receptor blockers, LDL: low density lipoprotein cholesterol, HDL: high density lipoprotein cholesterol, SD: standard deviation, SMD: standard mean difference.
