## Supplementary Table 7 for "Metformin use is associated with lower risks of dementia, anxiety and depression: The Hong Kong Diabetes Study"

Supplementary Table 7. Baseline and clinical characteristics of patients with type 2 diabetes mellitus who died or remained alive on follow-up before and after 1:1 propensity score matching.

* for SMD≥0.2

|  | **Before matching** |  |  | **After 1:1 matching** |  |  |
| --- | --- | --- | --- | --- | --- | --- |
| **Characteristics** | **All-cause mortality (N=34287) Mean(SD);N or Count(%)** | **Alive (N=55484)**  **Mean(SD);N or Count(%)** | **SMD** | **All-cause mortality (N=18373) Mean(SD);N or Count(%)** | **Alive (N=24115)**  **Mean(SD);N or Count(%)** | **SMD** |
| ***Demographics*** |  |  |  |  |  |  |
| Male gender | 16280(47.48%) | 24678(44.47%) | 0.06 | 9085(49.44%) | 11069(45.90%) | 0.07 |
| Female gender | 18007(52.51%) | 30806(55.52%) | 0.06 | 9288(50.55%) | 13046(54.09%) | 0.07 |
| Baseline age, years | 73.6(10.5);n=34287 | 62.0(11.6);n=55484 | 1.05* | 74.2(10.7);n=18373 | 62.1(12.0);n=24115 | 1.07* |
| <50 | 1025(2.98%) | 8341(15.03%) | 0.43* | 523(2.84%) | 3820(15.84%) | 0.46* |
| [50, 60) | 2836(8.27%) | 15438(27.82%) | 0.53* | 1468(7.98%) | 6401(26.54%) | 0.51* |
| [60, 70) | 6155(17.95%) | 15962(28.76%) | 0.26* | 3006(16.36%) | 6685(27.72%) | 0.28* |
| [70, 80) | 13918(40.59%) | 13006(23.44%) | 0.37* | 7289(39.67%) | 5819(24.13%) | 0.34* |
| >=80 | 10353(30.19%) | 2737(4.93%) | 0.70* | 6087(33.13%) | 1390(5.76%) | 0.74* |
| ***Past comorbidities*** |  |  |  |  |  |  |
| CAIDE score | 4.8(1.2);n=34287 | 4.0(1.6);n=55484 | 0.56* | 4.9(1.2);n=18373 | 4.1(1.7);n=24115 | 0.57* |
| CHA-DS-VASc  score | 3.0(1.6);n=34287 | 1.7(1.4);n=55484 | 0.86* | 3.1(1.6);n=18373 | 1.7(1.4);n=24115 | 0.89* |
| Charlson comorbidity index | 4.1(2.0);n=34287 | 2.3(1.7);n=55484 | 0.96* | 4.3(2.1);n=18373 | 2.4(1.8);n=24115 | 1.0* |
| Number of comorbidities | 2.3(2.3);n=34287 | 1.1(1.6);n=55484 | 0.57* | 2.6(2.4);n=18373 | 1.2(1.7);n=24115 | 0.66* |
| Hypertension | 13398(39.07%) | 11236(20.25%) | 0.42* | 8467(46.08%) | 6033(25.01%) | 0.45* |
| Heart failure | 2915(8.50%) | 1192(2.14%) | 0.29* | 1906(10.37%) | 633(2.62%) | 0.32* |
| Coronary heart disease | 4432(12.92%) | 3647(6.57%) | 0.22* | 3174(17.27%) | 2179(9.03%) | 0.25* |

| Atrial fibrillation | 1843(5.37%) | 882(1.58%) | 0.21* | 1164(6.33%) | 471(1.95%) | 0.22* |
| --- | --- | --- | --- | --- | --- | --- |
| Hemiplegia or paraplegia | 854(2.49%) | 488(0.87%) | 0.13 | 542(2.94%) | 250(1.03%) | 0.14 |
| Renal diseases | 2192(6.39%) | 863(1.55%) | 0.25* | 1499(8.15%) | 523(2.16%) | 0.27* |
| Neurologic | 532(1.55%) | 194(0.34%) | 0.12 | 496(2.69%) | 153(0.63%) | 0.16 |
| Ophthalmic | 1456(4.24%) | 731(1.31%) | 0.18 | 1305(7.10%) | 533(2.21%) | 0.23* |
| Liver diseases | 242(0.70%) | 98(0.17%) | 0.08 | 170(0.92%) | 75(0.31%) | 0.08 |
| Ventricular tachycardia/fibril lation | 378(1.10%) | 179(0.32%) | 0.09 | 233(1.26%) | 102(0.42%) | 0.09 |
| Anemia | 3723(10.85%) | 1331(2.39%) | 0.35* | 3414(18.58%) | 1021(4.23%) | 0.46* |
| AMI | 775(2.26%) | 554(0.99%) | 0.1 | 398(2.16%) | 269(1.11%) | 0.08 |
| COPD | 1262(3.68%) | 778(1.40%) | 0.15 | 778(4.23%) | 408(1.69%) | 0.15 |
| IHD | 3880(11.31%) | 3147(5.67%) | 0.20* | 2438(13.26%) | 1561(6.47%) | 0.23* |
| PVD | 654(1.90%) | 260(0.46%) | 0.13 | 436(2.37%) | 163(0.67%) | 0.14 |
| Stroke/TIA | 3058(8.91%) | 2178(3.92%) | 0.20* | 1858(10.11%) | 1102(4.56%) | 0.21* |
| Gastrointestinal bleeding | 1449(4.22%) | 1026(1.84%) | 0.14 | 962(5.23%) | 594(2.46%) | 0.14 |
| Obesity | 382(1.11%) | 701(1.26%) | 0.01 | 162(0.88%) | 268(1.11%) | 0.02 |
| ***Medications*** |  |  |  |  |  |  |
| Metformin v.s. sulphonylurea | 24575(71.67%) | 43952(79.21%) | 0.18 | 8661(47.13%) | 12583(52.17%) | 0.1 |
| Other anti- diabetic drugs | 9037(26.35%) | 16083(28.98%) | 0.06 | 5523(30.06%) | 6738(27.94%) | 0.05 |
| ACEI/ARB | 18805(54.84%) | 25462(45.89%) | 0.18 | 9666(52.60%) | 9791(40.60%) | 0.24* |
| Beta blockers | 13330(38.87%) | 17712(31.92%) | 0.15 | 7358(40.04%) | 8319(34.49%) | 0.11 |
| Calcium channel blockers | 18492(53.93%) | 20022(36.08%) | 0.36* | 9763(53.13%) | 8655(35.89%) | 0.35* |
| Diuretics | 9185(26.78%) | 8214(14.80%) | 0.30* | 5721(31.13%) | 4265(17.68%) | 0.32* |
| Stains | 9311(27.15%) | 13049(23.51%) | 0.08 | 5391(29.34%) | 5899(24.46%) | 0.11 |

| Nitrates | 214(0.62%) | 69(0.12%) | 0.08 | 198(1.07%) | 58(0.24%) | 0.1 |
| --- | --- | --- | --- | --- | --- | --- |
| Sulfadiazine | 23494(68.52%) | 37145(66.94%) | 0.03 | 9954(54.17%) | 11558(47.92%) | 0.13 |
| Antihypertensive drugs | 279(0.81%) | 89(0.16%) | 0.09 | 260(1.41%) | 80(0.33%) | 0.12 |
| Lipid-lowering drugs | 431(1.25%) | 254(0.45%) | 0.09 | 384(2.09%) | 204(0.84%) | 0.1 |
| Anticoagulants | 64(0.18%) | 15(0.02%) | 0.05 | 64(0.34%) | 15(0.06%) | 0.06 |
| Antiplatelets | 430(1.25%) | 154(0.27%) | 0.11 | 380(2.06%) | 132(0.54%) | 0.13 |
| ***Laboratory tests*** |  |  |  |  |  |  |
| Lymphocyte, x10^9/L | 1.7(1.7);n=5853 | 2.0(0.8);n=4092 | 0.21* | 1.7(1.7);n=4901 | 1.9(0.8);n=2410 | 0.18 |
| Neutrophil, x10^9/L | 5.8(2.8);n=5853 | 5.1(2.4);n=4092 | 0.26* | 5.8(2.8);n=4901 | 5.2(2.5);n=2410 | 0.24* |
| Platelet, x10^9/L | 256.2(91.8);n=2727 | 262.6(77.9);n=2995 | 0.07 | 251.7(89.2);n=1292 | 261.4(75.1);n=1200 | 0.12 |
| Potassium, mmol/L | 4.3(0.5);n=21029 | 4.3(0.4);n=29372 | 0.01 | 4.2(0.5);n=11498 | 4.2(0.5);n=11761 | 0.04 |
| Albumin, g/L | 36.9(5.6);n=8399 | 40.4(4.2);n=8205 | 0.7* | 36.6(5.7);n=6091 | 40.2(4.4);n=4120 | 0.72* |
| Sodium, mmol/L | 139.1(3.5);n=21041 | 139.5(2.9);n=29387 | 0.12 | 139.0(3.5);n=11508 | 139.6(3.0);n=11769 | 0.19 |
| Urea, mmol/L | 8.1(5.1);n=19003 | 5.8(2.4);n=26027 | 0.57* | 9.2(6.2);n=10666 | 6.1(3.0);n=10567 | 0.64* |
| Creatinine, umol/L | 129.0(134.4);n=19122 | 84.2(48.4);n=26155 | 0.44* | 155.8(172.3);n=10726 | 90.5(70.8);n=10630 | 0.5* |
| Total protein, g/L | 73.6(7.4);n=8363 | 75.2(6.0);n=8189 | 0.24* | 73.4(7.7);n=6063 | 75.1(6.3);n=4110 | 0.25* |
| Alkaline phosphatase, U/L | 83.2(44.8);n=18196 | 74.0(24.6);n=23754 | 0.25* | 86.9(46.3);n=10197 | 76.3(26.8);n=9865 | 0.28* |
| Aspartate transaminase,  U/L | 34.2(88.2);n=1571 | 29.7(46.4);n=1736 | 0.06 | 33.9(67.4);n=911 | 32.6(59.0);n=800 | 0.02 |
| Alanine transamise, U/L | 22.6(20.7);n=15321 | 27.3(24.8);n=18174 | 0.2* | 23.0(20.5);n=8696 | 28.4(25.5);n=7845 | 0.23* |

| Bilirubin, umol/L | 13.5(14.0);n=1150 | 13.3(6.8);n=840 | 0.02 | 13.5(14.0);n=1150 | 13.3(6.8);n=840 | 0.02 |
| --- | --- | --- | --- | --- | --- | --- |
| Triglyceride, mmol/L | 1.7(1.4);n=24952 | 1.7(1.4);n=40104 | 0.01 | 1.7(1.4);n=13688 | 1.8(1.4);n=17142 | 0.04 |
| Low-density lipoprotein, mmol/L | 2.9(0.9);n=15263 | 2.9(0.8);n=25829 | 0.11 | 2.8(0.9);n=8323 | 3.0(0.9);n=10877 | 0.17 |
| LDL calculated, mmol/L | 2.8(0.9);n=17389 | 2.9(0.9);n=28863 | 0.15 | 2.7(0.9);n=9351 | 2.9(0.9);n=11724 | 0.19 |
| Total cholesterol, mmol/L | 4.7(1.1);n=25039 | 4.9(1.0);n=40167 | 0.13 | 4.7(1.1);n=13741 | 5.0(1.0);n=17176 | 0.2 |
| High-density lipoprotein, mmol/L | 1.2(0.4);n=21867 | 1.2(0.3);n=35782 | 0.07 | 1.2(0.4);n=11848 | 1.2(0.3);n=14729 | 0.1 |
| HbA1c, g/dL | 11.9(2.0);n=6331 | 13.2(1.7);n=4906 | 0.7* | 11.7(2.0);n=5260 | 13.0(1.8);n=2888 | 0.7* |
| Glucose, mmol/L | 9.2(4.6);n=10239 | 8.7(4.0);n=19100 | 0.12 | 8.9(4.5);n=7913 | 8.2(3.8);n=12938 | 0.18 |

CAIDE: cardiovascular risk factors, aging, and incidence of dementia, AMI: acute myocardial infarction, COPD: chronic obstructive pulmonary disease, IHD: ischemic heart disease, PVD: peripheral vascular disease, TIA: transient ischemic attack, ACEI: angiotensin-converting-enzyme inhibitors, ARB: angiotensin II receptor blockers, LDL: low density lipoprotein cholesterol, HDL: high density lipoprotein cholesterol.
