## Supplementary Table 8 for "Metformin use is associated with lower risks of dementia, anxiety and depression: The Hong Kong Diabetes Study"

Supplementary Table 5. Adjusted HRs (and 95% CIs) for comparing metformin and sulphonylurea users using cause-specific or subdistribution hazard models before and after 1:1 propensity score matching.

* for p≤ 0.05, ** for p ≤ 0.01, *** for p ≤ 0.001

| **Model** | **Outcome** | **Metformin v.s. sulphonylurea (Before matching)**  **HR [95% CI]** | **P value** | **Metformin v.s. sulphonylurea (After 1:1 matching)**  **HR [95% CI]** | **P value** |
| --- | --- | --- | --- | --- | --- |
| Cause-specific hazard model | New-onset dementia | 0.88[0.82, 0.96] | 0.0025** | 0.77[0.62, 0.89] | 0.0014** |
|  | New-onset anxiety disorder and depression | 0.67[0.60, 0.75] | <0.0001*** | 0.59[0.51, 0.69] | <0.0001*** |
|  | All-cause mortality | 0.68[0.67, 0.70] | <0.0001*** | 0.81[0.78, 0.83] | <0.0001*** |
| Subdistribution hazard model | New-onset dementia | 0.89[0.77, 0.97] | 0.0003** | 0.82[0.69, 0.91] | 0.0004*** |
|  | New-onset anxiety disorder and depression | 0.69[0.57, 0.78] | <0.0001*** | 0.62[0.57, 0.75] | <0.0001*** |
|  | All-cause mortality | 0.71[0.66, 0.82] | <0.0001*** | 0.83[0.76, 0.92] | <0.0001*** |

HR: hazard ratio, CI: confidence interval.
