## Supplementary Table 9 for "Metformin use is associated with lower risks of dementia, anxiety and depression: The Hong Kong Diabetes Study"

Supplementary Table 6. Risks of new diagnosis of dementia, anxiety disorder/depression and all-cause mortality in matched cohorts associated with for metformin vs. sulphonylurea users with multiple approaches using the propensity score.

* for p≤ 0.05, ** for p ≤ 0.01, *** for p ≤ 0.001

| **Outcome** | **HR after PS stratification [95% CI];P value** | **HR after HDPS matching [95% CI];P value** | **HR after PS IPTW [95% CI]; P value** |
| --- | --- | --- | --- |
| New-onset dementia | 0.81[0.72-0.97];<0.0001*** | 0.85[0.75-0.95];<0.0001*** | 0.85[0.73-0.97];<0.0001*** |
| New-onset anxiety or depression | 0.71[0.58-0.85];<0.0001*** | 0.68[0.51-0.82];<0.0001*** | 0.7[0.65-0.85];<0.0001*** |
| All-cause mortality | 0.75[0.67-0.86];<0.0001*** | 0.75[0.71-0.87];<0.0001*** | 0.74[0.71-0.86];<0.0001*** |

HR: hazard ratio; CI: confidence interval; PS: propensity score, HDPS: high dimensional propensity score, IPTW: inverse probability of treatment weighting.
