## Supplementary Figure 1 for "Metformin use is associated with lower risks of dementia, anxiety and depression: The Hong Kong Diabetes Study"

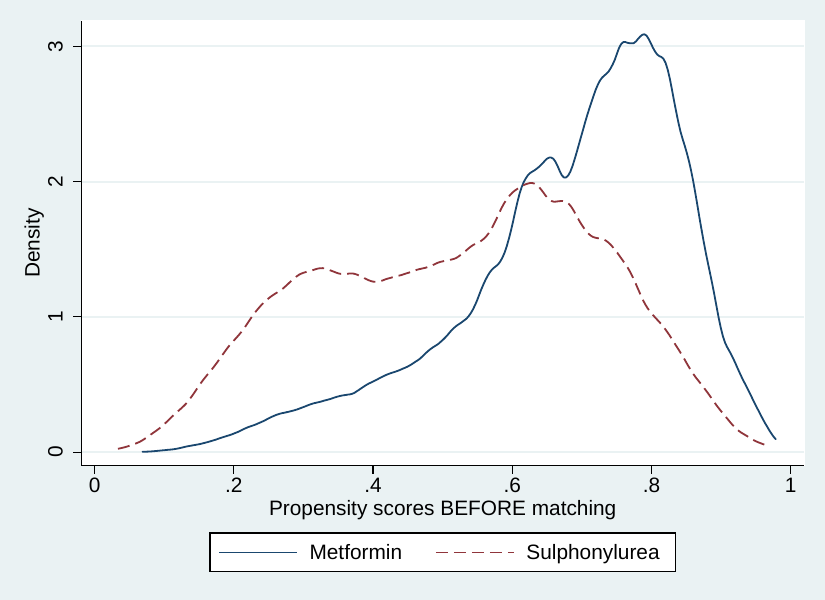

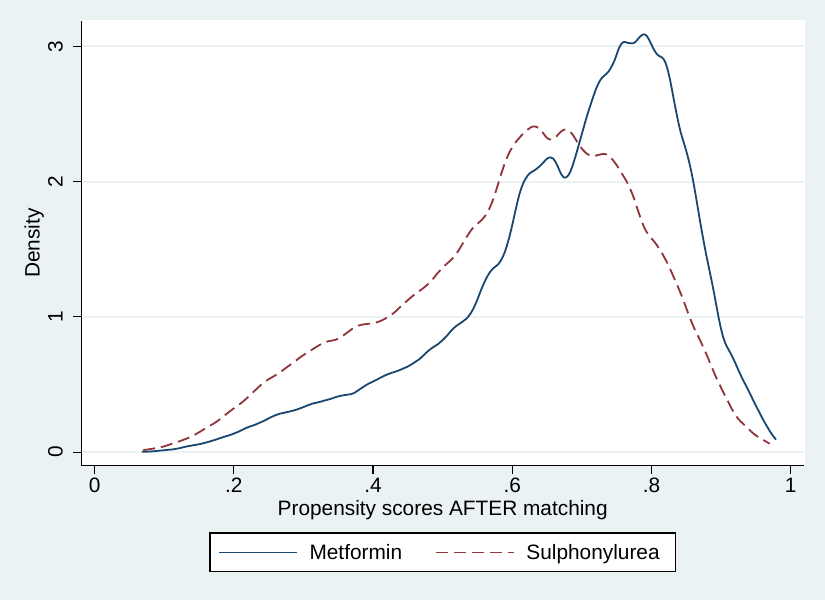

**Supplementary Figure 1. Density as a function of propensity score using the nearest neighbor search strategy.**
